# Suicide during the COVID-19 pandemic in Japan

**DOI:** 10.1101/2020.08.30.20184168

**Authors:** Takanao Tanaka, Shohei Okamoto

## Abstract

There is an emerging concern that the COVID-19 pandemic could harm psychological health and exacerbate suicide risk. Utilising month-level suicide records covering the entire Japanese population in 1,848 administrative units, we assessed whether suicide mortality changed during the pandemic. Employing difference-in-difference estimation, we found that monthly suicide rates declined by 14% during the initial five months (February to June 2020), which could be due to a number of complex reasons, including the government’s generous subsidies, reduced working hours, and school closure. In contrast, monthly suicide rates increased by 16% in the second wave (July to October 2020), with the magnitude greater among females (37%) and children and adolescents (49%). The adverse impact of the COVID-19 pandemic may remain, while its modifiers (e.g. government subsidies) may not be sustained for long. Hence, effective suicide prevention, particularly among vulnerable populations, should be an important public health consideration.

## Main

The COVID-19 pandemic has affected every aspect of life. As the virus has spread globally^1^, anxious individuals have voluntarily engaged in physical distancing and reduced their economic activities to prevent infection. To contain the virus, governments have implemented large-scale costly interventions in an unprecedented fashion: citizens and communities are requested to limit social contacts, avoid social gatherings, close schools, and stop unnecessary business activities. Thus far, most scientific and clinical attention has been given to identifying the disease’s direct physical risk^2,3^ and its prevention^4-6^. However, the end of this pandemic is seemingly nowhere near. This raises an emerging concern in a different public health arena: the pandemic could adversely affect people’s mental health^7,8^ and, in a more pressing scenario, suicide fatalities could increase^9,10^.

While suicide is rarely due to a single factor, and reasons for changes to suicide prevalence are extremely complex, prior literature suggests that the pandemic may affect the suicide rate in various ways. Together with fear, uneasiness, and anxiety caused by the threat of the disease, social distancing can lead to impaired social and family relationships; increased loneliness, boredom, and inactivity; and restricted access to healthcare services, potentially inducing mental illness and elevated suicidal behaviours.^10,11^ Financial insecurity and loss of employment are well-known risk factors for suicide^12,13^. Thus, the pandemic-driven economic recession could increase potential suicidal deaths^14^. Not surprisingly, existing studies suggest that past epidemics such as the Spanish Flu and Severe Acute Respiratory Syndrome (SARS) led to increased suicide rates.^15,16^ At the same time, the current pandemic might have reduced some of the stress from workplaces and social interactions (such as commuting, and school bullying), and government financial support could have partially alleviated the pandemic’s adverse impacts. However, given the unprecedented magnitude, ubiquity, and complexity of the ongoing public health crisis, adequate preventive measures to reduce the risk of suicide will be required. To formulate effective policy responses, policymakers, healthcare professionals, and researchers need a credible assessment of suicide prevalence during this pandemic.

However, reliable empirical evidence regarding the link between the COVID-19 pandemic and suicide mortality remains scarce. An inclusive assessment requires harmonised data that cover representative and sufficiently large samples but are collected at a disaggregated level^17,18^. Such data also should include information from both the pre-COVID-19 period (to serve as the baseline samples) and the COVID-19 period. However, existing studies use readily available and convenient data that could easily generate biased insights; many studies rely on some measures of suicidality rather than suicide mortality^19-24^ and most of them compare suicidal behaviours using snapshot data during the pandemic without pre-pandemic baseline samples^19,20,22,23^. Even when studies use real suicide mortality, some rely on data that cover non-representative sub-samples^25-27^, while others compare the whole suicide or suicidality trend before and during the pandemic, which might capture common time trend, seasonality, or temporal time shocks across individuals^25-31^ (we discuss why these time-series analysis and before-after comparison can be problematic in the Method section).

In this study, we provide large-scale evidence linking the COVID-19 outbreak to suicide fatalities using a city-by-month level dataset covering the entire Japanese population, more than 120 million people (Fig. 1; see Supplementary Note 1 and Supplementary Table 1 for the details of data). Since the confirmation of the first case in Japan, the nation has been hit by two large-scale COVID-19 outbreaks. In response, the national and local governments implemented various preventive interventions, such as nationwide school closure and state of emergency, and local business restrictions. Although the government concurrently provided generous financial support for citizens and enterprises, accounting for as much as 10% of Japan’s annual GDP^32^, people’s lives were still profoundly affected; for instance, the geographic mobility had dropped below pre-pandemic level and the unemployment rate has increased for nine consecutive months (Supplementary Fig. 1 and Supplementary Note 2). Since the Japanese suicide rate is the seventh-highest among high-income countries^33^ and has been among the top ten causes of death for the last two decades,^34^ there have been rising concerns that the COVID-19 crisis may increase suicide deaths.

**Fig. 1.**
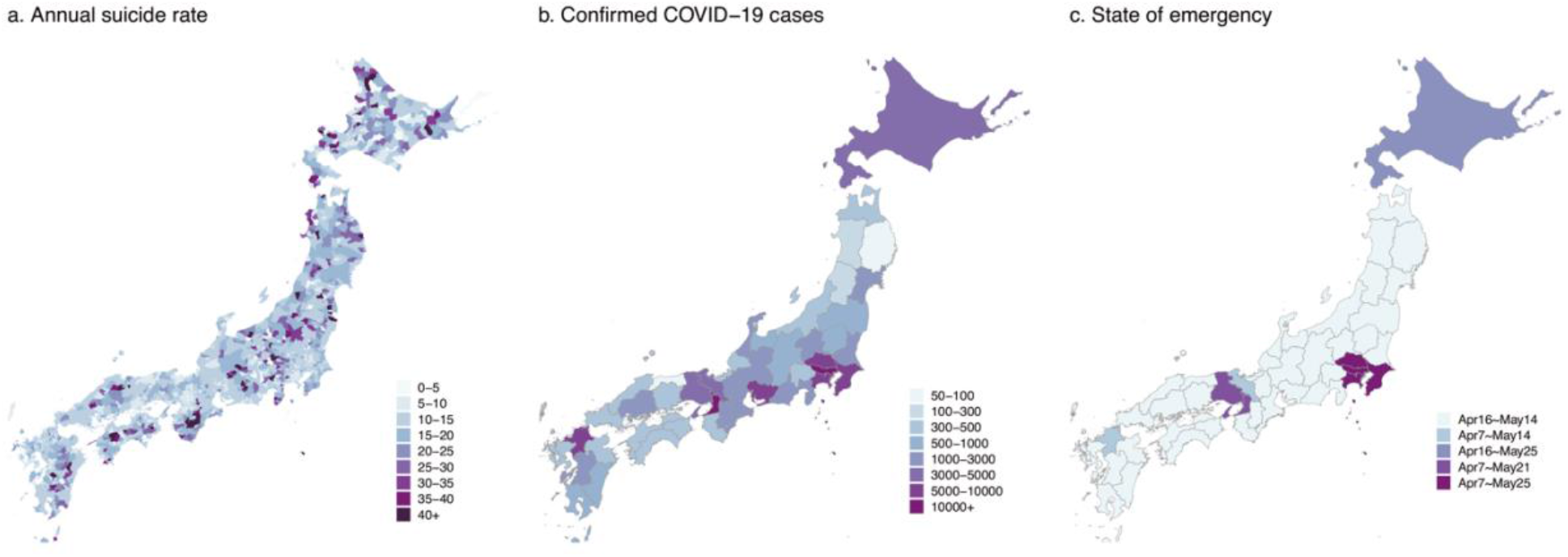
Distribution of suicide rate, COVID-19, and health interventions in Japan: **Panel a** describes the mean number of suicides in a month in each city during the period of our study. **Panel b** shows the number of confirmed COVID-19 cases on October 31 in each prefecture. **Panel c** presents the length of the state of emergency, which was implemented and lifted depending on the severity of the disease outbreak. These maps are created by the authors using the shape file. See Supplementary Note 1 for the detail.

Our data have some notable advantages for assessing whether suicide mortality changed during the pandemic and the subsequent health interventions. First, our data cover both pre-pandemic and pandemic-era samples from November 2016 to October 2020, so that we can investigate the relative change in suicide rate compared to the pre-pandemic baseline. (We describe the suicide trend in Extended Data Figs. 1 and 2.) In particular, to construct a reliable control condition (without pandemic), we employ a difference-in-differences (DID) estimation. We assess whether suicide rates during the pandemic varied compared to the corresponding seasons in the previous years, after controlling for the variation in the overall suicide level across years (long-term suicide trend). Moreover, our data are collected from 1,848 administrative units, which enables us to control for various confounding factors. Specifically, our regression includes two sets of high-dimensional fixed effects: a city-by-month fixed effects control for month-specific shocks in each city (e.g. seasonality in suicide rate, monthly local events, or climatic conditions), and a city-by-year fixed effects control for a year-specific shock in each city (e.g. macroeconomic trend, industrial or population structural changes, or suicide trends). (We describe details in the Methods section.)

The effects of the pandemic might not be evenly distributed across populations. To identify the vulnerable populations, we further analyse heterogeneous impacts across gender and age groups. Historically, male adults have faced the highest suicide risk in Japan. During the pre-pandemic period in our dataset, the male suicide rate is about 2.3 times as high as that of females (per-month suicide rate is 18.1 per million for males and 7.8 per million for females from November 2016 to January 2020, see Supplementary Table 1), and is even higher among male adults (21.8 per million among males aged 20-69). Because previous literature suggests financial and economic distress could trigger suicide fatalities particularly among males^35^, and the COVID-19 pandemic brought about non-negligible disruptions in the labour market, we might expect its adverse impact to be greater among such populations. (We summarise the suicide trends in Japan in Supplementary Note 3.) Alternatively, older adults face a heightened risk of infection and death from COVID-19^2,3^, which could amplify their fear and distress about virus transmission. Additionally, existing studies suggest that, unlike normal economic recessions, this pandemic has gendered impacts: social distancing disproportionately affects female-dominant employment^36^, and stay-home orders increase household tasks, and even domestic violence, which could disproportionately impair well-being among females^37^. These suggest that the effects of the COVID-19 pandemic could be intensified among these previously lower-risk populations.

Two analyses complement our empirical analyses. First, we investigate whether the impacts vary across job status, which can indicate through which channels the pandemic and health interventions can affect suicide mortality. For instance, we might expect the government subsidies to mitigate the adverse impacts differently across individuals (such as employed, retired, and unemployed) and firm owners (self-employed). Further, household dynamics (increase in housework, domestic violence) might disproportionately affect housewives’ psychological health, and unusual school calendars (nationwide school closure, and the reductions in outdoor activities and social connections) might affect students’ mental well-being. Lastly, we assess whether the effect of the pandemic varied across different types of cities using the dimensions of original suicide risks, pandemic associated risks (COVID-19 prevalence, health interventions, and economic downturns), and base socio-economic status (income and urban populations). For example, we investigate whether the suicide trend varied between cities with high and low per-capita income.

## Results

### Main Results

The suicide rate substantially declined during the first wave of the COVID-19 pandemic (February to June 2020), but rapidly increased during the second outbreak (July to October 2020) (Fig. 2a). The DID estimates during the first outbreak, adjusting for permanent unobserved city-by-year and city-by-month determinants of the suicide rate, show that the overall suicide rate declined by 14% (incidence rate-ratios [IRR]: 0.86, 95% confidence interval [CI]: 0.82 to 0.90) compared to the same season in previous years. In contrast, it increased by 16% (IRR: 1.16, 95% CI: 1.11 to 1.21) amid the second outbreak (Fig. 2a). During the second wave, an increasing suicide trend has been apparent, and the suicide rate increased by 38% in October 2020 (IRR: 1.38, 95% CI: 1.27 to 1.49; the full results are in Supplementary Tables 2 and 3).

**Fig. 2.**
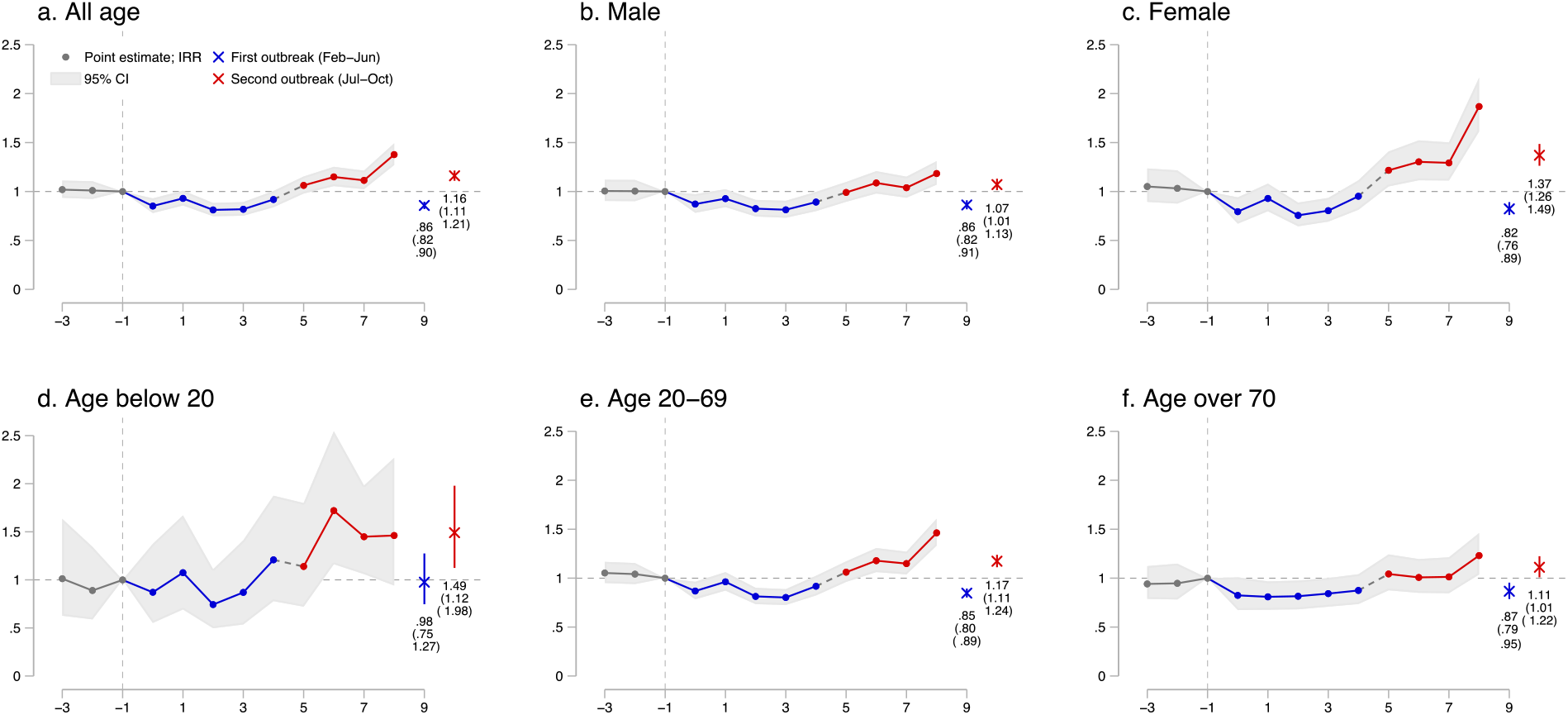
The effects of COVID-19 pandemic on suicide across gender and age groups using DID and event-study approaches: **Panel a** describes the results of the event study using all the pooled samples. **Panels b and c** show the results using suicide rates among males and females. **Panels d, e, and f** present the results on different age groups. In all graphs, the lines represent the point estimates coloured by grey before the pandemic, by blue during the first outbreak of the COVID-19, and by red during the second outbreak, with shaded areas showing the 95% confidence interval. The blue and red circle and line denote the DID result and its 95% confidence interval during the first and second outbreaks, respectively. The full results are described in Supplementary Tables 2-4. All regressions include city-by-year fixed effects and city-by-month fixed effects and are weighted by the population, and standard errors are clustered at the city level. N= 61,209 (a), 54,583 (b), 35,712 (c), 1,896 (d), 53,164 (e), and 34,703 (f). The separated observations are dropped (see Methods), and suicide data of children and adolescents are aggregated at prefectural level.

Using the event-study approach, from estimated coefficients, we confirm that the assumption for parallel trends is not violated during the pre-treatment period, as average suicide rates in 2016-2019 and 2020 are not different (Fig. 2: k <= −1) (Supplementary Table 4). These results are robust to the inclusion of the time-varying weather variables and prefecture-specific quadratic time trends (Supplementary Fig. 2a). Moreover, the adoption of the OLS model, instead of the Poisson model, does not affect our main findings (Supplementary Fig. 2b).

To ascertain that the results are not driven by either the common time trends or common year-specific shocks across cities or regions, we conduct a placebo test. Extended Data Fig. 3a shows that when using the placebo samples, the estimated coefficients are close to zero, and our real estimate for the first outbreak is much smaller than the lower bound of the coefficients’ 95% CI (IRR: 0.96 to 1.04), while the estimate for the second outbreak is much larger than the upper bound of the coefficients’ 95% CI (IRR: 0.96 to 1.04). This analysis suggests that our results are not driven by a spurious correlation. These patterns hold when we conduct the event-study regression analogously using the placebo samples, validating the strong relationship between the pandemic and the suicide rate. (Extended Data Fig. 3b)

On average, 1,596 individuals died by suicide in each month during our study period (November 2016-October 2020). Our back-of-the-envelope calculation finds that there were 1,074 averted suicide deaths during the initial outbreak (95% CI: 830 to 1,488) from February to June 2020 but there were 970 additional suicide deaths during the subsequent outbreak (95% CI: 630 to 1,170) from July to October 2020 (Extended Data Fig. 4). In the corresponding period, the number of direct deaths from COVID-19 was 1,765 (as of October 31), suggesting that the magnitude of the suicidal effects is not negligible.

### Heterogeneity Across Gender and Age Groups

We investigate the heterogeneous impacts of the pandemic across gender, age groups, and periods. We find some noteworthy variation in the magnitude of the pandemic effects (full results in Supplementary Table 5).

First, during the nationwide state of emergency, suicide reduction among adults (individuals aged 20-69 years) was particularly salient (Fig. 3a). In the middle of the first wave, to slow down viral transmission, the national government declared a state of emergency, where individuals were requested to stop unnecessary business, reduce social contacts, and stay home if possible. During this period, suicide deaths among adults declined by about 21∼27% (Fig. 3a, male adults, IRR: 0.79, 95% CI: 0.73 to 0.85; female adults, IRR: 0.73, 95% CI: 0.65 to 0.83), and this is about twice as much as the decline during other periods in the first wave. In contrast, the reduction in the suicide rate among the individuals aged 70 and over was not remarkably larger during the state of emergency (Figs. 3a and 3b).

**Fig. 3.**
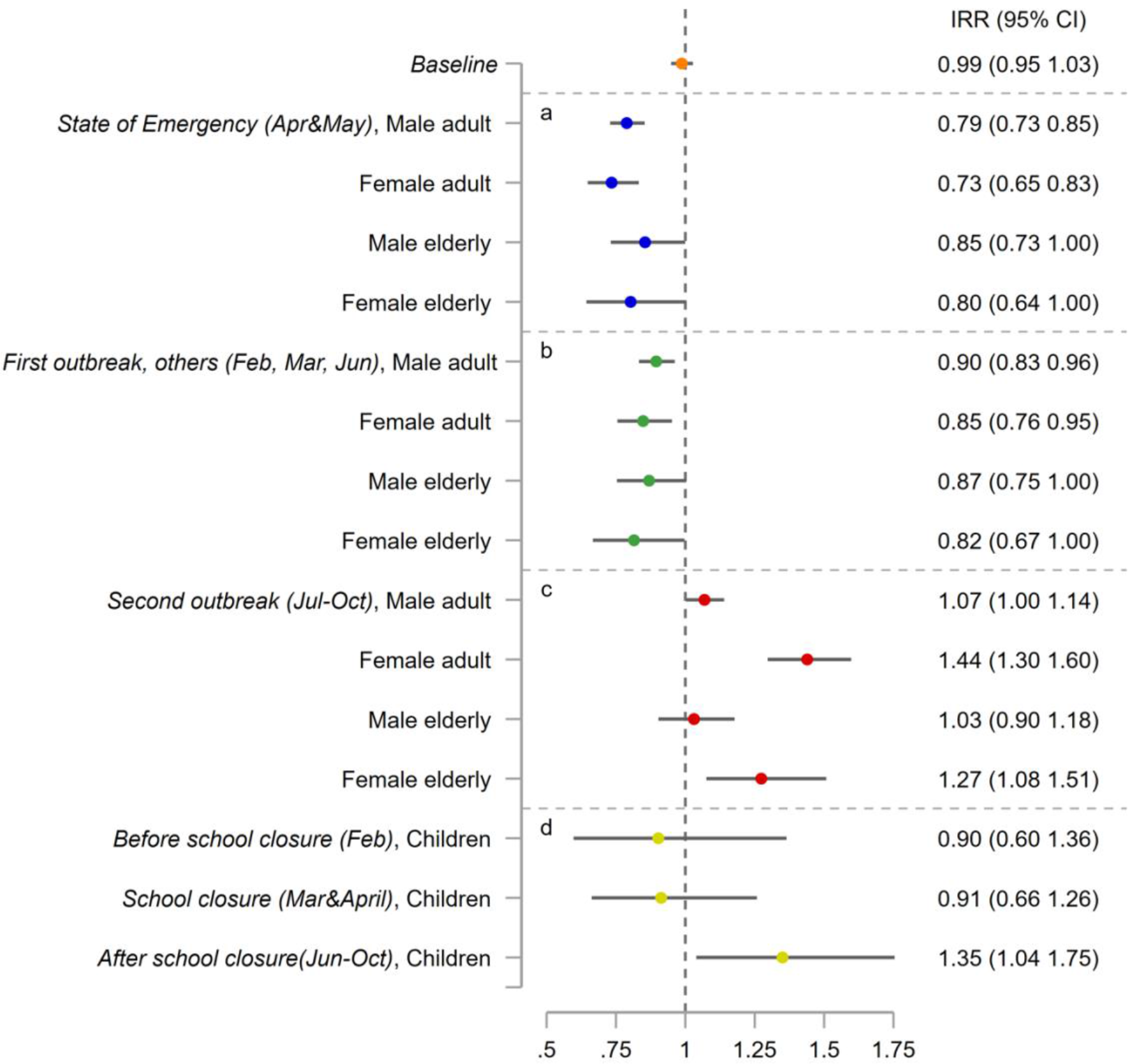
Heterogeneous effects of the COVID-19 pandemic among age groups and gender, before and after the stage of emergency, and the school closure: Here, we assign April and May as the period of the state of emergency (SOE), February-June as the first outbreak period, and July-October as the second outbreak period. Baseline is a result when all samples are pooled. The circle and line denote the DID result and its 95% confidence interval in each period, respectively. **Panel a** represents the heterogeneous effects among age groups and gender during the state of emergency (SOE). **Panels b and c** describe heterogeneity across age and gender groups during the first outbreak (except for SOE) and second outbreak, respectively. **Panel d** shows the effects on students during the school closure (March and April) and other periods. The full results are represented in Supplementary Table 5. All regressions include city-by-year fixed effects and city-by-month fixed effects and are weighted by the population, and standard errors are clustered at the city level. N= 61,209 (all), 47,317 (male adults), 26,319 (female adults), 24,478 (male elderly), 16,531 (female elderly), and 1,896 (children and adolescents). The separated observations are dropped (see Methods), and suicide data of children and adolescents are aggregated at prefectural level.

Second, the increase in suicide fatalities in the second wave is primarily driven by females and children and adolescents (individuals aged below 20 years) (Figs. 2 and 3). Suicide mortality increased by 37% among females (Fig. 2c, IRR: 1.37, 95% CI: 1.26 to 1.49), and this is about 5 times greater than males (Fig. 2b, IRR: 1.07, 95% CI: 1.00 to 1.14). In October 2020, its increase was particularly substantial; we observe that suicide rate among females increased by 82% (Supplementary Table 3, IRR: 1.82, 95% CI: 1.62-2.04). The suicide rate among children and adolescents was also heightened in the second wave, which mostly corresponds to the period after the end of the nationwide school closure (Fig. 2d,IRR: 1.49, 95% CI: 1.12 to 1.98).

### Heterogeneity Across Job Status

The suicide rate decreased among those employed, retired, and unemployed in the first wave of the epidemic, but mostly increased in the second wave (Fig. 4a-Fig. 4c). These patterns are identical to the population as a whole. In contrast, we observe some dissimilar suicide trends among specific sub-populations.

**Fig. 4.**
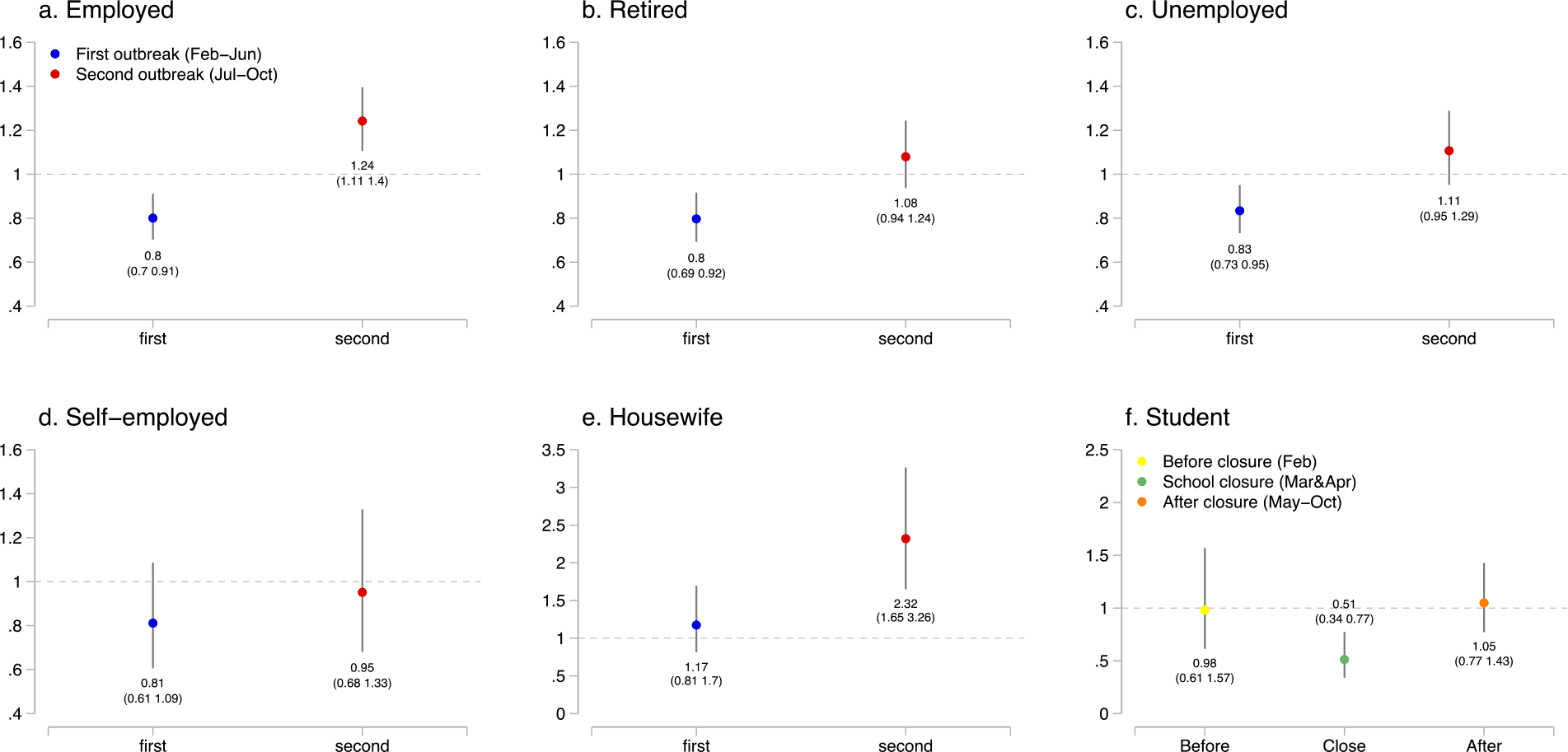
Heterogeneous effects of the COVID-19 pandemic among job status: This graph describes the results of the effects of the pandemic on suicide across those with different job status during the first and second outbreaks of COVID-19, and during school closure for students. The blue and red circle and line denote the DID result and its 95% confidence interval during the first and second outbreaks, respectively. For each panel, we use different outcomes. We use the number of the suicide rate among those whose jobs are classified as either employed (**panel a**), retired (**b**), unemployed (**c**), self-employed(**d**), housewife (**e**), or student (**f**). The full results are represented in Supplementary Table 6. All regressions include city-by-year fixed effects and city-by-month fixed effects and are weighted by the population, and standard errors are clustered at the city level. N= 10,723 (a), 9,146 (b), 9,246 (c), 4,124 (d), 3,854 (e), and 3,220 (f). The separated observations are dropped (see Methods). Also see Supplementary Note 1 for the data used for this analysis.

We find that the suicide rate among the self-employed did not decline in either the first or second wave (Fig. 4d). In contrast, suicides among housewives (defined as adult women who are married and not employed in wage labour) increased during all pandemic periods (Fig. 4e). In particular, from July to October 2020, the rate rose by 132% (IRR: 2.32, 95% CI: 1.65 to 3.26). We also find that suicide deaths by students declined by about 49% during the school closure (Fig. 4f, IRR: 0.51, 95% CI: 0.34 to 0.77) (full results in Supplementary Table 6). The results for students are slightly different than those for children and adolescents. We describe these data in Supplementary Note 1.

### Heterogeneity Across Cities

In Fig. 5, we investigate whether the effect of the pandemic varied across different types of cities (full results in Supplementary Table 7). First, we compare cities with higher suicide risks (if the suicide rate before the pandemic is high, the city is classified as a high-suicide-risk city) with cities with lower suicide risks (Fig. 5a). We might expect suicide effects during the pandemic to be intensified in the high-risk cities. However, the results show the opposite; only cities which previously had low suicide rates saw an increase during the pandemic.

**Fig. 5.**
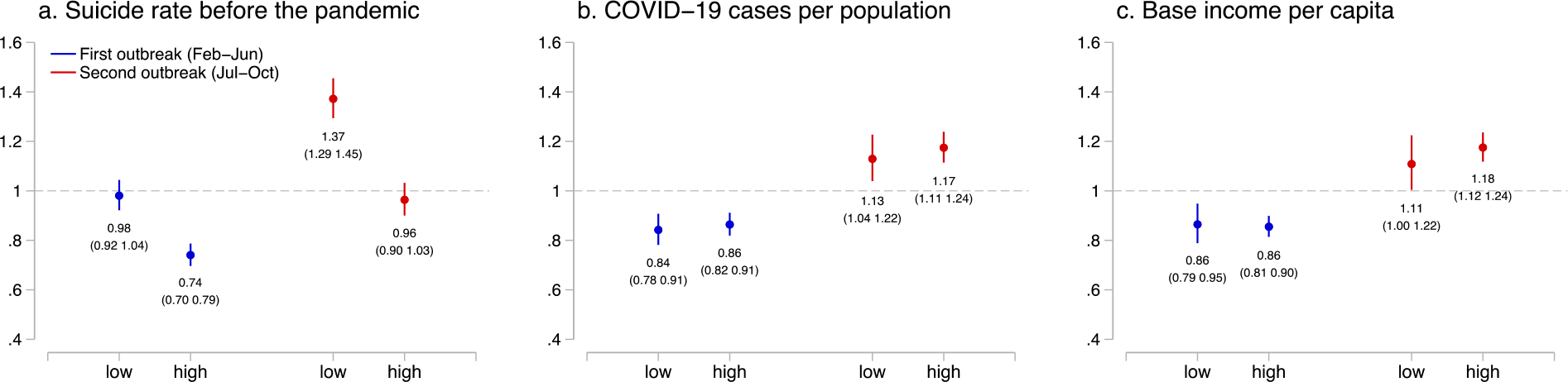
Heterogenous effects of the COVID-19 pandemic across geography: This graph describes the results of the effects of the pandemic on suicide during the first and second outbreaks across locations with different original suicide risk before the pandemic (**panel a**), prefectural level COVID-19 confirmed cases (**b**), and base income per capita (**c**). The blue and red circle and line denote the DID result and its 95% confidence interval during the first and second outbreaks, respectively. If the variable in a city or a prefecture is lower than the median, the city or the prefecture is defined as “low”. The full results are represented in Supplementary Table 7. All regressions include city-by-year fixed effects and city-by-month fixed effects and are weighted by the population, and standard errors are clustered at the city level. N= 30,134 (a, low), 31,075 (a, high) 24,246 (b, low), 36,963 (b, high), 26,166 (c, low), and 35,043 (c, high). The separated observations are dropped (see Methods).

We also investigated heterogeneity with respect to disease associated risks. We used COVID-19 prevalence (measured by total confirmed cases per million population in October 2020) and find the effects do not vary across cities regarding the dimensions (Fig 5b). We also utilised variation in intensity of the health intervention (Extended Data Fig. 5a: measured by Google mobility change at workplaces from January 2020 to after February 2020), and economic shocks (Extended Data Fig. 5b: measured by the changes in unemployment from October-December 2019 to April-September 2020), and did not find notable disparities in the impact. Note that, because these data are available at prefectural level, but not at city level, these results should be interpreted with caution.

Lastly, we find the effects are not heterogeneous in terms of the base socio-economic status, such as income per capita (Fig 5c: measured in 2018) and urban population (Extended Data Fig. 5c: measured in 2018).

## Discussion

Utilising high-frequency data that cover the entire Japanese population, we investigate whether suicide mortality changed during the pandemic. In the Methods section, we argue that the use of time-series analysis^25-27,29-31,38^ and simple before-after comparison^26,27,29,30,38^ could easily generate biased estimates. Instead, we use a DID model with high-dimensional fixed effects that allow us to control for a number of potential confounders. Consistent with emerging concerns, we find that suicide deaths increased from July to October 2020 (IRR: 1.16, 95% CI: 1.11 to 1.21), although the effects appeared several months after the pandemic started. Notably, an increasing suicide trend has been apparent during this period.

The COVID-19 pandemic has affected every aspect of life. In Japan, the unemployment rate has increased for nine consecutive months since the epidemic was recognised (Supplementary Fig. 1c). Social interactions and mobility also have been substantially restricted by individual choice or by government intervention (see mobility index in Supplementary Fig. 1d). Fear and anxiety regarding infection are persistent. Given these circumstances, while the reasons behind suicide are extremely complex, there are a number of factors that could explain the rapid increase in the suicide rate during the second COVID-19 outbreak.

We find that the effects of the pandemic are not evenly distributed across populations. Compared to the historical suicide pattern, we find that this time is different. First, the previous suicide rate among males was 2.3 times higher than that among females (from November 2016 to January 2020, see Supplementary Table 1), and the suicide increase among males after earlier financial crises was larger than that among females^39^. In contrast, during the second wave of the COVID-19 pandemic, the increase in the suicide rate among females (IRR: 1.37, 95% CI: 1.26 to 1.49) was about five times greater than that among males (IRR: 1.07, 95% CI: 1.01 to 1.13), and that among housewives was even more so (IRR: 2.32, 95% CI: 1.65 to 3.26). (The suicide level among males remained higher, but disparities decreased.) These results are consistent with recent studies that find that this crisis unevenly affects female-dominant industries^36^, and stay-home orders magnify the working mother’s burden^40^. When we look at the Japanese labour market, we also find similar patterns: the decrease in female employment is more pronounced than in male employment (Fig. 6a), and non-regular workers (56% of females and 22% of males are non-regular workers) are unevenly affected during this pandemic (Fig. 6b). In addition, domestic violence, mostly harming women (more than 95% of all cases), increased (Fig. 6c, see Supplementary Note 1 for data). All of this could have harmed women’s psychological health.

**Fig. 6.**
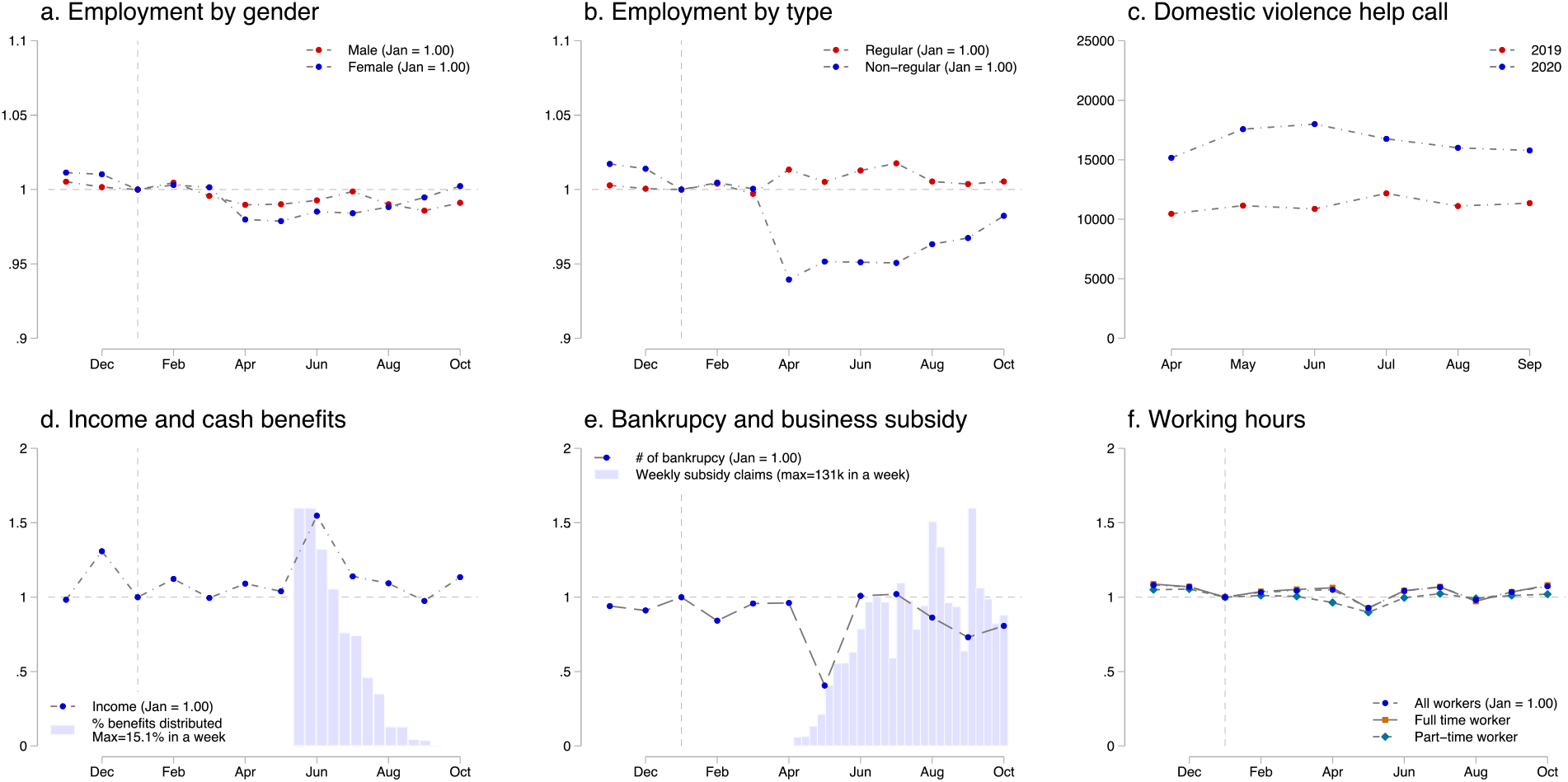
Mechanisms of the pandemic affecting suicide rate: **Panel a** shows employment by gender, **Panel b** shows the employment rate by types of employment (regular and non-regular workers). **Panel c** reports the number of calls for help related to domestic violence from April to December in 2019 and 2020. The data from October to March are not available. **Panel d** represents the average household income, after the bonus payment was extracted. The bar graph shows the share of the cash benefits (100,000 = 940USD) distributed in each week. **Panel e** depicts the trend of bankruptcies and the claims for business subsidy. **Panel f** displays the average working hours for full-time workers and part-time workers before and during the pandemic. In a, b, d, e, and f, data are standardized so that the value takes one month before the pandemic (January 2020).

The suicide rate among children and adolescents also increased in the second wave of the pandemic (IRR: 1.49, 95% CI: 1.12 to 1.98). This may be because the pandemic also excessively affects younger workers, who are more likely to be low-skilled, and employed on less secure work contracts. In fact, the decline in the employment rate during the disease outbreak is greater among those generations. Furthermore, the second outbreak corresponds to the period when schools (elementary to high school) were reopened after the nationwide school closure. Previous work reports that schooling could be a risk factor for violence^41^ and suicide^42^ among students. After a few months of life without any school activities during the pandemic, the stress from returning to school could have been exacerbated. These factors might have amplified children’s and adolescents’ psychological depression.

Immediately after the COVID-19 outbreak (February to June 2020), however, we find a notable reduction in suicide rates (IRR: 0.86, 95% CI: 0.82 to 0.90). The finding might have been unexpected, yet is consistent with the emerging studies and statistics which find that suicide deaths decreased in Norway^30^, the UK^43^, Germany^27^ and Peru^44^, and did not change in Greece^31^, Massachusetts (US)^25^, Victoria (Australia)^45^, and China (outside Wuhan)^46^ (Suicide deaths in Nepal increased^29^), when these countries were placed under strict lockdowns. Also, existing studies often report a drop followed by a delayed increase in suicide rates after national disasters, including Hurricane Katrina in 2005 or the 9/11 terrorist attack in 2001. (The initial decline is called the pulling-together effect or honeymoon effect^47-49^; see Supplementary Note 4.) Hence, suicide decline in the initial phase of this public health crisis may not be surprising.

In Japanese contexts, several additional mechanisms could explain this decline in the initial stage. In response to the crisis, the national government provided several subsidies and benefits to citizens and enterprises (Supplementary Note 1 and 2), which might have reduced economic distress. We observe that about 80% of the cash benefits was distributed to citizens before June (all citizens were eligible to receive cash benefits of 100,000 yen ≈ 940 USD), and suicide among individuals (employed, unemployed, and retired) decreased only before June (Fig. 4a-c). Additionally, claims for business subsidies grew rapidly after May until October, and the number of bankruptcies remained lower than the pre-pandemic level (Fig. 6e). The suicide rate among the self-employed did not increase during the periods.

We also see that working hours for both full-time and part-time workers declined substantially (10%∼20%) from April to May (Fig. 6f). Overwork and commuting are well-recognised risk factors for suicide, particularly among the working population in Japan^50^; hence, reduced working hours and work-from-home policies could have improved people’s productivity, life satisfaction and mental health^51-53^. This explanation is consistent with our finding of the largest decline in suicide rates among adults during the state of emergency (male adult, IRR: 0.79, 95% CI: 0.73 to 0.85, female adult, IRR: 0.73, 95% CI: 0.65 to 0.83), which mainly comprise the working population.

Finally, school closure may have reduced psychological burdens on children and adolescents, which could have resulted in suicide prevention. We have found that the suicide rate declined substantially among students during the nationwide school closure, March and April 2020 (IRR: 0.51, 95% CI: 0.34 to 0.77).

Our results offer a number of important insights on suicide mortality during the pandemic that might remain relevant even after normal life resumes. First and foremost, the suicide trend in Japan could remain escalated in the long term. In the absence of effective pharmaceutical interventions (e.g. a vaccine or antiviral treatments), pandemic-related suicidal risk factors (e.g. disease recurrence, social distancing, and economic downturns) would remain. While we have argued that massive government subsidies and benefits might have might have contributed to the prevention of suicides in the beginning of the epidemic, such generous financial support might not be sustainable for long. Hence, society has to monitor the overall suicide trends, so that it can immediately consider to take policy responses.

In addition, our results suggest that, to formulate effective prevention strategies amid the COVID-19 pandemic, customised approaches, rather than conventional ones, are needed. We have found that, unlike normal economic circumstances, this pandemic disproportionately affects the psychological health of children and adolescents, and females (especially housewives). Additionally, we find that only cities which previously had low suicide rates saw an increase suicide in deaths during the pandemic. Therefore, the prevention strategy might need to target these vulnerable populations and locations.

Our results could also suggest directions to prevent suicide even after normal life resumes. We argued that reduced working hours might have contributed to a reduction of suicide among adults during the state of emergency (April to May), and school closure might have had a protective role among students. These highlight the need to identify the exact factors associated with workplaces and school sessions which could affect the psychological well-being of working-populations and students.

We conclude by discussing the limitations of our study. First, while we have proposed potential mechanisms for the linkage between the pandemic and suicides, it is challenging to disentangle the contributions of each factor. To do so, one needs variation in the timing and intensity of each contributor (e.g. disease prevalence, government interventions, economic shocks, financial support, and working conditions). However, the COVID-19 pandemic affects almost every community and citizen concurrently, and our analyses could not fully utilise such variation.

Moreover, we could not investigate the effect among specific subgroups of interest. For example, mental health consequences among healthcare professionals have been of great concern as they are taking on extraordinary burdens during the pandemic^54,55^. Similarly, suicide effects among those with a high risk of case fatalities (those with the presence of comorbidities)^2,3^, financial strain (low-skilled or low-income workers)^36^, or social isolation (including those who need mental health care or those with cognitive disorders) should be carefully monitored.

Additionally, suicide data may not be precise during the COVID-19 pandemic. It is possible that some suicide deaths were not found, or were reported after delays. Also, suicide might have been misclassified as a different cause of death because the pandemic could have disrupted the reporting process. If any of these is the case, our estimates could be understated.

Finally, we want to emphasise that the results of our study may not apply to other communities or countries because our study is founded on the unique Japanese public health, economic, cultural, and social contexts. Particularly, during our study period, the confirmed COVID-19 cases per population in Japan were only 2.9% of those in the US and 12.7% of those in Germany (Supplementary Fig. 3a). Moreover, the Japanese government’s health intervention is among the most lenient, based on “request” rather than “enforcement,” ensuring a high degree of individual freedom (Supplementary Fig. 3b). Nevertheless, the national government provided very generous fiscal support for households and enterprises, accounting for 10% of the annual GDP (Supplementary Fig. 3c). These together could make a difference in the extent of the epidemic’s impact on mental well-being. Therefore, to protect global psychological health by preventing escalated suicide rates, continuous assessment of the effect of the pandemic in each society is crucial.

## Methods

### Data

We use city-by-month data on suicide records from November 2016 to October 2020, covering all the suicide deaths among a total of 126 million citizens in Japan. The data are derived from suicide statistics published by the Ministry of Health, Labour, and Welfare, and they include information such as the number of suicides by age, gender, employment status, site, and day of the week^56,57^. The dataset includes 76,626 (monthly average 1,596) suicides in 1,848 cities (N = 88,512) with the average monthly suicide rate at 12.8 per million population. Males account for 68.2% of total suicides, and, in particular, male adults (between 20 and 69 years) contribute to about half of this total (50.2%) (Supplementary Table 1). Using these data, we assess how suicide rates vary before and during the pandemic.

We also use other datasets to supplement our analysis, including the number of COVID-19 infections, weather conditions, and macroeconomic conditions (i.e., bankruptcy, unemployment rates, consumption index, and diffusion index). Details for these data are available in Supplementary Note 1.

### Difference-in-Differences model

A central empirical challenge to estimating the effect of COVID-19 on suicide rates is to disentangle the effect of the pandemic from the long-term suicide trend and its seasonality. On average, the suicide rate has declined by 6.4% from 2017 to 2019 (Extended Data Fig. 2; if data are extended, it declined by about 25% from 2013 to 2019). In addition, before the pandemic, the average suicide rate in February is 5.1% higher than that in January. These imply that study design based on the before-after comparison could be problematic; if it compares the suicide levels before and during the COVID-19 outbreak^29^ (such as interrupted time-series analysis or regression discontinuity design), the estimates might capture the seasonal trend (particularly between January and February); alternatively, if it compares the suicide level relative to past years in the same season^26,27,30,38^, the estimate might be confounded by a long-term ascending or descending suicide trend. (In Supplementary Note 5 and Supplementary Fig. 4, we discuss why interrupted time-series analysis is problematic in our research context.)

The suicide trend and its seasonality also have to be accounted for at a disaggregated level, because they vary widely across locations (Extended Data Fig. 2). For instance, most regions had a declining suicide trend (Extended Data Fig. 2b1, and c1), although it increased in some regions (Extended Data Fig. 2d1). Relatedly, we observe that the suicide rate in summer is higher in some locations (Extended Data Fig. 2b2), while that pattern is reversed in others (Extended Data Fig. 2e2). If we eliminate such location-specific trends and seasonality (we regress suicide rate on city-by-month, and city-by-year fixed effects, and eliminate those effects), suicide patterns seem to be different from the observed trends (Extended Data Fig. 2a3∼e3). These make the point that the time-series analysis, which compares national suicide trends, could easily generate biased estimates^25-27,29-31,38^. Instead, accurate estimation requires a quasi-experimental research design and harmonised data, by defining reasonable location-specific control conditions (counterfactual without the pandemic).

By leveraging our disaggregated but comprehensive dataset, we adopt the Difference-in-Differences (DID) estimation with high-dimensional fixed effects. Our model is designed to overcome the empirical challenges. First, the model compared the difference in suicide rates before (November to January) and during the virus outbreaks (February to October) with the difference in the corresponding period in the previous three years (November 2016 to October 2019). Because the model focuses on the relative difference before and during the sudden pandemic within a year, the overall suicide level across years (long-term suicide trend) will be cancelled out. Second, we include city-by-month fixed effect and city-by-year fixed effect. These rich sets of fixed effects allow us to isolate the pandemic effects from the location-specific suicide trend and seasonality.

In particular, we specify the following model;

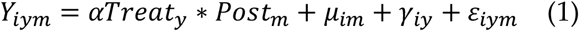

where *Y* denotes suicide rates in city *i* in month *m* in year *y* (a year includes 12 months from November to October), and *α* is the parameter of interest, which denotes the impacts of the COVID-19 pandemic on suicide rates. *Post*_*m*_ is a binary variable that takes 1 if periods of observations corresponded to months between February (when the COVID-19 outbreak became salient and the national government launched the nationwide anti-contagion policies) and October. It takes 0 if periods correspond to months from November to January regarded as the “pre-treatment” period prior to the COVID-19 pandemic. *Treat*_*y*_ takes 1 if the year is 2020 (November 2019 to October 2020) and 0 otherwise (November 2016 to October 2019). In this model, suicide trends between February and October from 2016 to 2019 serve as a control condition (counterfactual), after accounting for the level across years, with the assumption that we encounter only common shocks between the control and the treatment periods (during the pandemic).

We include city-by-month fixed effect and city-by-year fixed effect denoted by *μ*_*im*_and *γ*_*iy*_. The former flexibly controls for month-specific shocks in each city, such as seasonality in the suicide rate, monthly local events, or climatic conditions^58,59^. The latter controls for year-specific shocks in each city, such as macroeconomic trends, industrial or population structural changes, or suicide trends.

Suicide trends during the pandemic could vary across periods depending on the size of the ongoing outbreak, people’s response, and the government’s health interventions. Specifically, Japan faced two large COVID-19 outbreaks (Supplementary Note 2), and we might expect the suicide trend to vary in each wave. Therefore, we estimate:

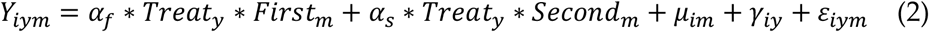

where *First*_*m*_ denotes the dummy variable, which takes 1 during months corresponding to the first outbreak (February to June), and *Second*_*m*_ takes 1 during months corresponding to the second outbreak (July to October). Similarly, we estimate how the suicide trends differ during the state of emergency (April and May 2020), and the school closure (March and April 2020).

Our outcome variables of interest, suicide rate, is left-skewed and non-negative. Specifically, 58.7% of the city-by-month suicide rate takes zero during our study period. Therefore, we use a Poisson-pseudo-maximum-likelihood (PPML) estimator to specify the equations (1) and (2)^60,61^. The adjusted model for equation (2) can be written as:

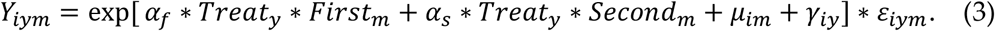

We use a package “ppmlhdfe” to estimate the regression, with options “weight”, “absorb” and “cluster” in STATA version 16 to implement all the Poisson regression analyses^60^. We report the estimated coefficient in the form of incidence rate-ratios (IRR). For this estimation, the necessary condition for the existence of the estimates is non-existence of the “separated” observations^61^. Therefore, such observations are dropped in the procedure. Note that because more than 95% of city-by-month suicide rate among children and adolescents takes zero, we aggregate the data to the prefectural level for this specific cohort.

We cluster standard errors at the city level to allow arbitrary correlation over time within the same city. Additionally, all the regressions are weighted by population in 2018 so that cities with larger populations are given greater weights. Intuitively, these weights help to estimate the impact of the event on an average person instead of on an average city.

### Event-study Approach

The assumption for the DID estimator to be valid is that the pandemic period (February to October) in 2020 and the same periods in 2016-2019 would have parallel trends in suicide rates in the absence of the pandemic. If this assumption were not satisfied, the estimated parameter would be biased because the results could be driven by systematic differences between the treatment and control groups rather than the event of interest. To assess whether the parallel trends assumption would be reasonable, we adopt the event study approach by fitting the following equation^37,62^:

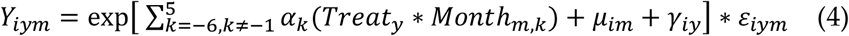

where *Month*_*m*_ takes 1 if the month corresponds to *k*, where *k* = −1 is set to be a month before the pandemic period (January). Intuitively, this casts the difference in suicide rates between 2020 season and 2016-2019 season in each month relative to *k* = −1: we expect the treatment group and control group to have a similar suicide rate before the disease outbreak becomes salient (*k* < 0), while we expect it to diverge after the outbreak (*k* ≥ 0).

### Heterogeneity

We estimate the heterogeneity impacts across different gender, age, job status, and geography. For age and gender, we re-estimate equation (3) by using suicide rate across gender and age groups (children and adolescents aged below 20 years, the working-age population aged 20-69 years, and older adults aged 70 years or over). For job status, we use suicide rate among employed, retired, unemployed, self-employed, housewife, and student.

For heterogeneous analysis across geography, we re-estimate equation (3) using the subsamples. We use city-level base suicide rate (measured from November 2016 to January 2020), base income per capita (measured in 2018), and base share of urban population (measured in 2018) to classify the samples, in that, if the variable in a city is above its median, the corresponding city is classified as a high-group. Similarly, we use the prefectural level COVID-19 prevalence (measured by total confirmed cases per million population in October2020), mobility restrictions (measured by Google mobility change at workplaces from January 2020 to after February 2020), and economic shocks (measured by the changes in unemployment from October-December 2019 to April-September 2020). We use prefectural level data for the sample classifications because the city-level data are not available. The details of these data are described in Supplementary Note 1.

### Placebo test

We perform a placebo test^37,63^ to ascertain that impacts of the pandemic on suicide rates are not driven by either common time trends or common shocks across different periods, using the following procedure. Using the data from November 2016 to October 2019, we randomly allocate treatment status to a year in the same period (February to June for the first outbreak, and July to October for the second outbreak) in each city and estimate the treatment effects analogously to equations (3) and (4). These equations can be written as:

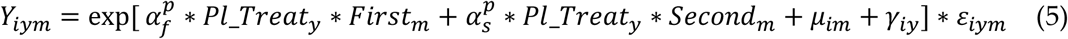

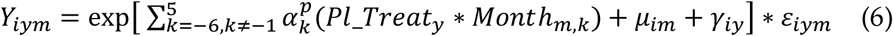

where *Pl*_*Treat*_*y*_ takes 1 if the treatment status is allocated in both the equations. Then, we compare the placebo results to the real estimates. We repeat these procedures 1,000 times. If there is an event causing higher suicide incidence in a specific region in a pre-pandemic period (e.g. cities in Tokyo prefecture in 2019 have unusually high suicide rates), our placebo results would include the spike in the estimated parameters. These results might imply that our main estimate is not driven by the disease outbreak, but by a random shock (or time trend) in some cities. We expect the placebo results (denoted by 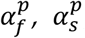, and 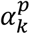) not to be statistically different from zero.

### Back-of-the-envelope calculation

To estimate the increased or decreased deaths from suicide during the pandemic, we estimate the following equation:

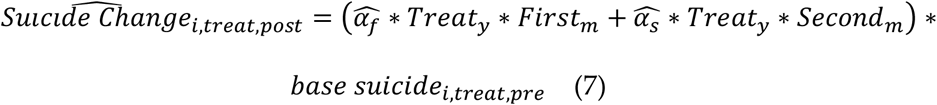

where 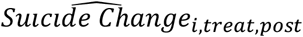 denotes the predicted suicide change in city *i* in treatment year (the year 2020) during the pandemic period (after February). This is computed by estimated coefficients (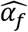 and 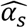) derived by specifying equation (3) for the period during the first outbreak (*Treat*_*y*_ ∗ *First*_*m*_, 5 months) and second outbreak (*Treat*_*y*_ ∗ *Second*_*m*_,4 *months*), and number of base suicide deaths (*base suicide*_*i,treat,pre*_). We then sum the changes in suicides in each city to compute change on the national scale.

## Data Availability

Data used in the paper are publicly available.

https://www.mhlw.go.jp/stf/seisakunitsuite/bunya/0000140901.html

https://github.com/sokamoto-github/Suicide-during-the-Covid-19-pandemic-in-Japan

## Data Availability

Data used in this paper are available at https://github.com/sokamoto-github/Suicide-during-the-Covid-19-pandemic-in-Japan.

## Code Availability

Code used in this paper are available at https://github.com/sokamoto-github/Suicide-during-the-Covid-19-pandemic-in-Japan.

## Acknowledgments

We thank Yuhang Pan and seminar participants at the 23^rd^ Tokyo Labour Economics Workshop for their insightful comments. TT thanks the Bai Xian Asia Institute for the scholarship support as a Bai Xian Scholar. SO is supported by the postdoctoral fellowship of the Japan Society for the Promotion of Science (No. 20J00394) and the Murata Science Foundation. These founders were not engaged in the conceptualisation, design, data collection, analysis, decision to publish, or preparation of the manuscript.

## Author Contributions

SO and TT conceptualised the study and carried out initial planning. TT retrieved and constructed the data set. TT carried out the statistical analysis, which was refined by SO for the final version. SO prepared the first draft of the report, which was revised by TT. All authors reviewed and contributed to the final draft and approved the final version for publication.

## Competing Interests

The authors declare no competing interests.

## Extended Data for

### Extended Data Figures

**Extended Data Fig. 1.**
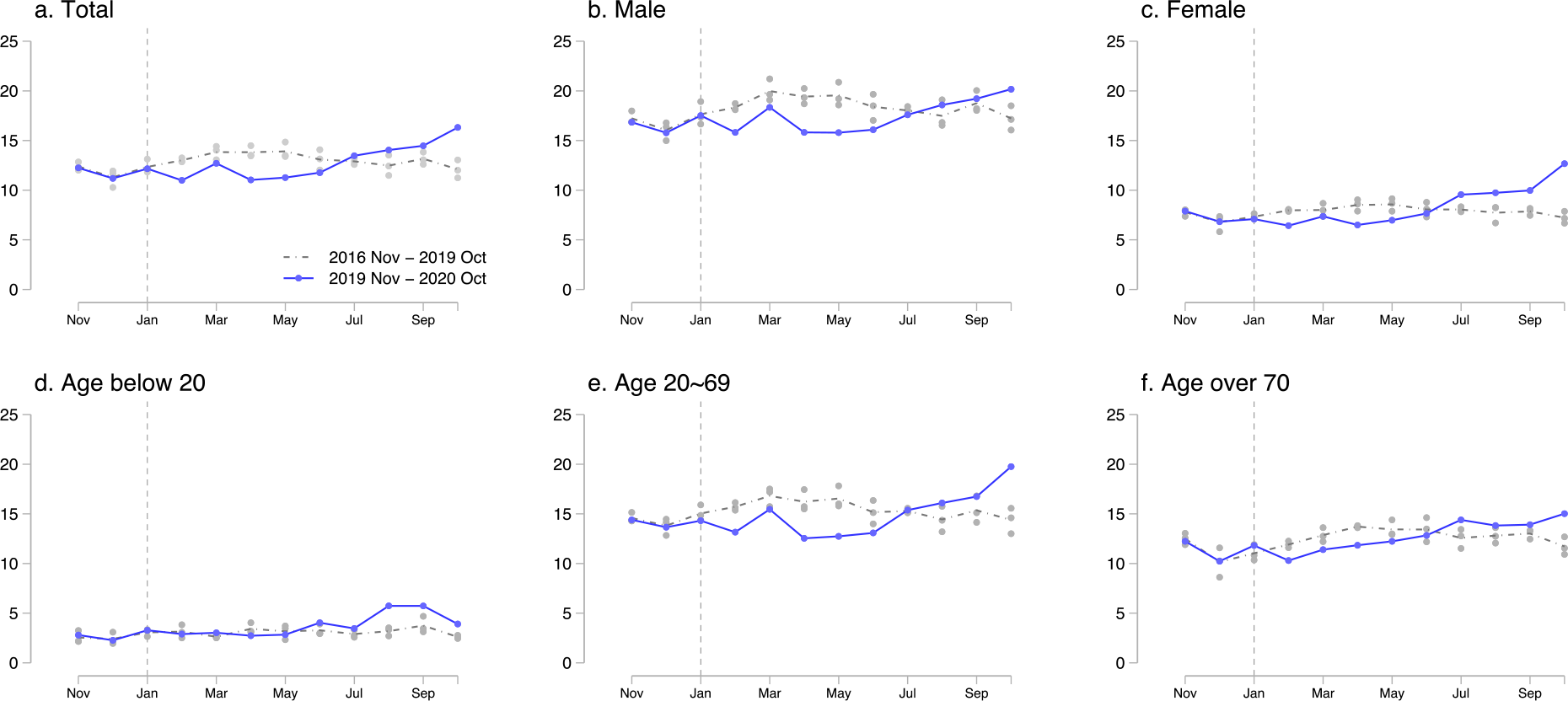
The trend of suicide across gender and age groups: **Panels a-f** show the trend of suicide from November 2016 to October 2020. The grey line represents the mean suicides rates for three years before the pandemic year (November 2016 to October 2019) with a grey circle denoting suicide each year. The light blue line shows the number of suicides from November 2019 to October 2020.

**Extended Data Fig. 2.**
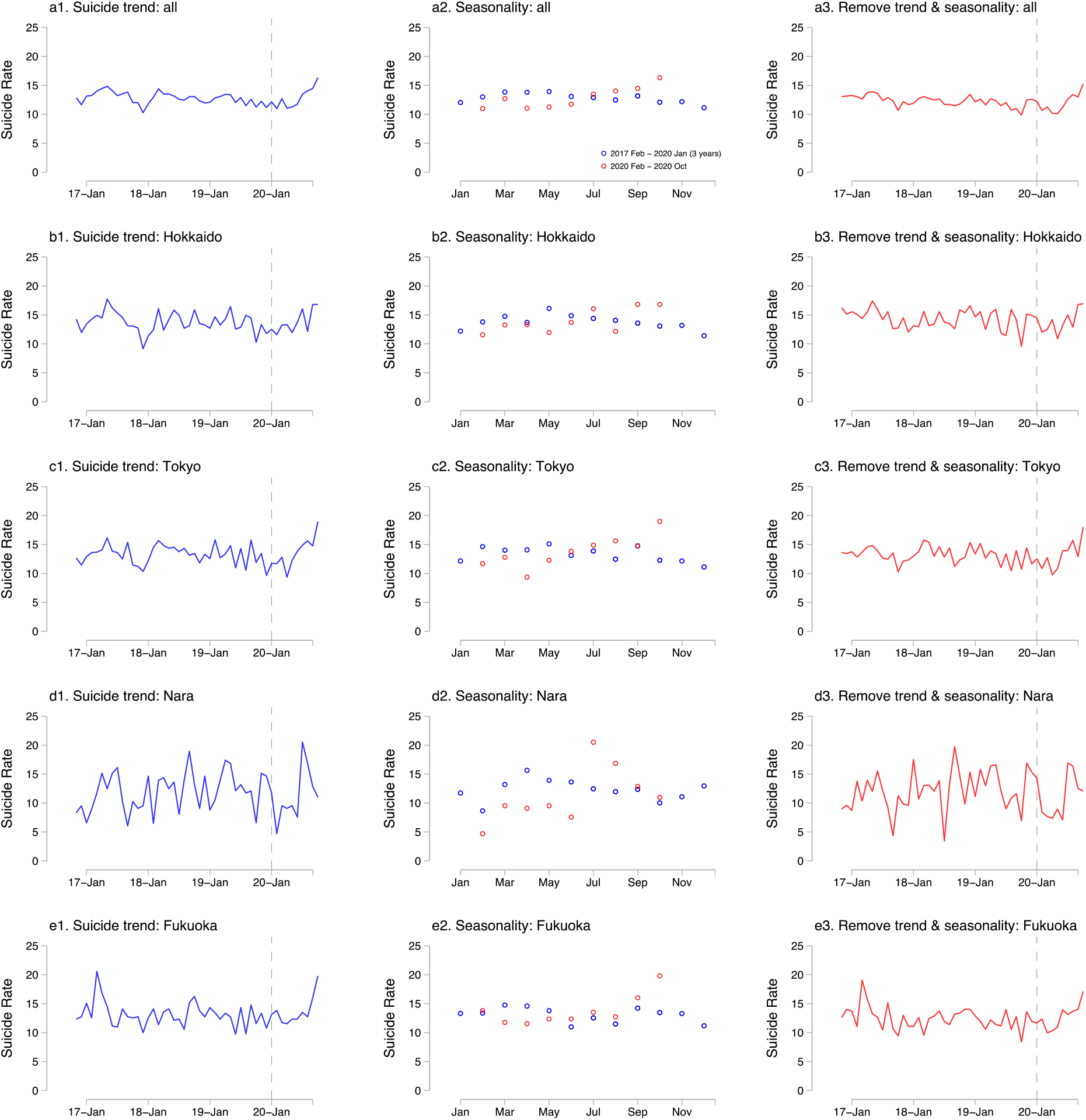
The trend and seasonality of suicide across regions: **Panels a1-e1** show the observed suicide rate in each prefecture. **Panels a2-e2** represent the seasonal suicide trend before the pandemic (Feb 2017-Jan 2020, three years) and during the pandemic. **Panels a3-e3** describe the suicide trends after eliminating the effects of suicide trend and seasonality. To do so, we regress suicide rate on city-by-month fixed effects, and city-specific suicide linear time trends, and eliminate these effects. **Panel a** uses all observations (1,848 cities), **Panel b** uses the cities in Hokkaido prefecture (177 cities), **Panel c** uses Tokyo prefecture (57 cities), **Panel d** uses Nara prefecture (39 cities), and **Panel e** uses Fukuoka prefecture (73 cities).

**Extended Data Fig. 3.**
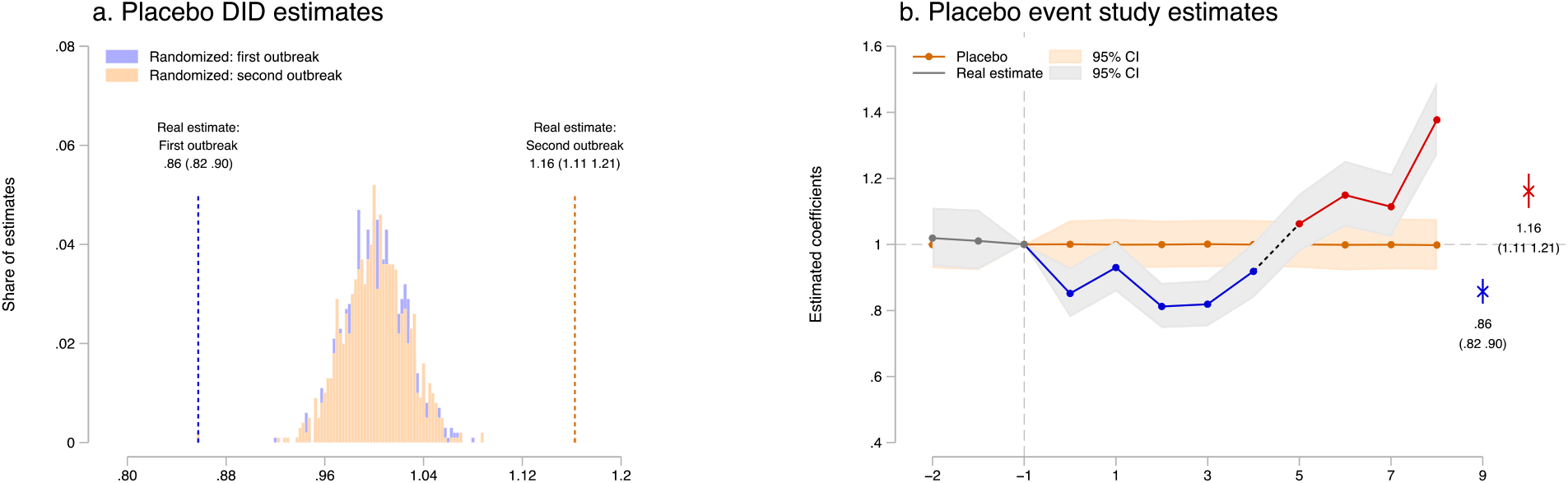
Robustness check: **Panel a** represents the results of the placebo test. Using samples from 2016 to 2019, we first randomly allocate treatment dummies within the city across the years. We then regress the suicide rate on the placebo treatment 1,000 times. The mean effect using the placebo sample is around zero, and the estimate of the first outbreak is much smaller than 95% CI of the placebo results and that of second outbreak is much larger than 95% CI of the placebo results. **Panel b** repeats event study regression using the placebo samples analogously. The estimated coefficient is close to zero in all periods. All regressions include city-by-year fixed effects and city-by-month fixed effects and are weighted by population, and standard errors are clustered at the city level. N= 61,209 (b). Each regression for 1,000 times has different number of observations (a). The separated observations are dropped (see Methods)

**Extended Data Fig. 4.**
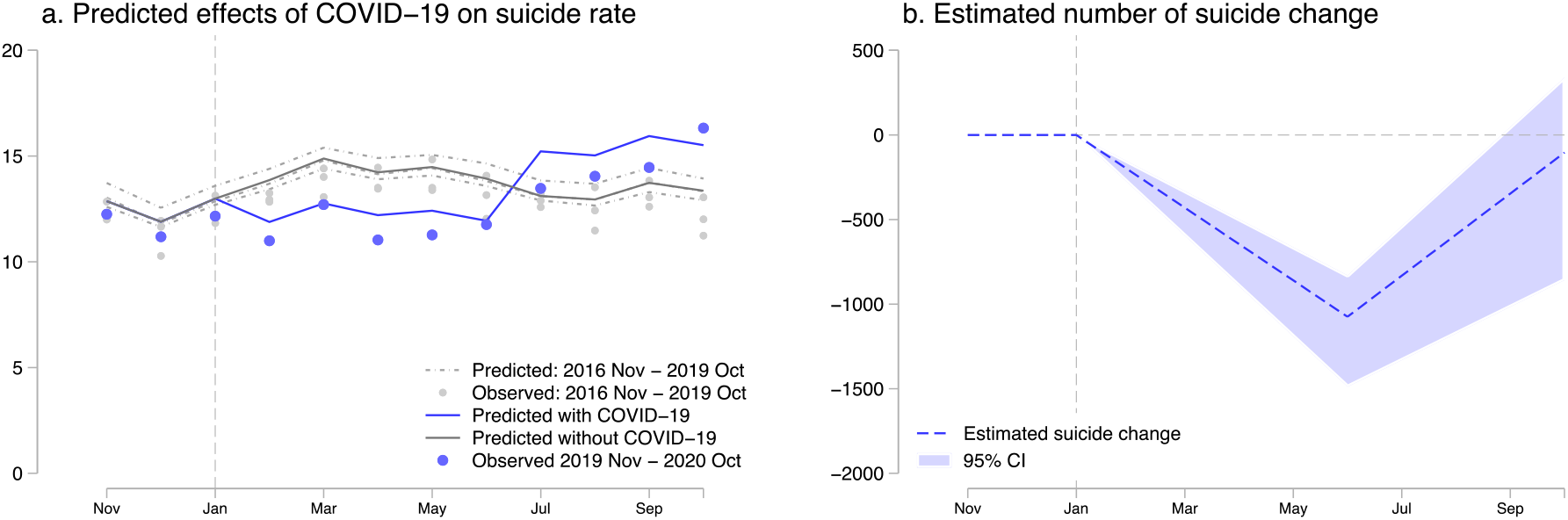
Back-of-the-envelope calculation for the predicted change in suicide deaths: **Panel a** describes the predicted change in suicide rate with COVID-19 and otherwise. The difference between the grey line (without COVID-19) and the blue line (with COVID-19) represents the effects of the pandemic. **Panel b** demonstrates the predicted increased or decreased deaths by suicide across periods. The blue dashed line represents the points estimate, while the light blue area shows the 95% CI. All regressions include city-by-year fixed effects and city-by-month fixed effects and are weighted by population, and standard errors are clustered at the city level. N= 61,209 (a). The separated observations are dropped (see Methods)

**Extended Data Fig. 5.**
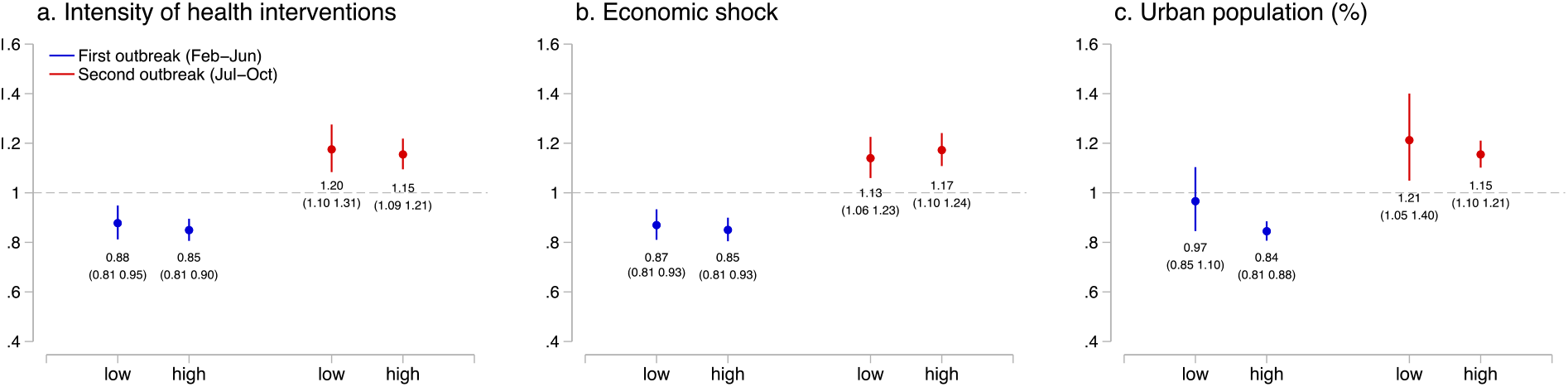
Additional analyses on heterogenous effects of the COVID-19 pandemic across geography: This graph describes the results of the effects of the pandemic on suicide during the first and second outbreaks across locations with different prefecture-level intensity of health interventions (**panel a**), prefecture-level economic shock (**b**), and base share of urban population (**c**). The intensity of health intervention is measured by the level of decline in the Google mobility index, and economic shock by the changes in unemployment rate. If the variable in a prefecture or city is lower than the median, the prefecture or city is defined as “low”. All regressions include city-by-year fixed effects and city-by-month fixed effects and are weighted by population, and standard errors are clustered at the city level. N= 23,530 (a, low), 37,679 (a, high), 26,004 (b, low), 35,205 (b, high), 18,784 (c, low), 42,425 (c, high). The separated observations are dropped (see Methods).

## Supplementary Materials for

### Supplementary Notes

#### Supplementary Note 1. Data sources and description

##### Map data of Japan (Fig. 1)

The shape file of Japan is obtained from the web page of ESRI Japan inc.^*1*^

The maps are created by authors.

##### COVID-19 cases in Japan (Fig. 1b)

The data are obtained from the web page of the Ministry of Health, Labour and Welfare in Japan^2^. The data include confirmed cases, deaths, and recovered at the prefectural level (47 prefectures in Japan).

##### Suicide rate among children and adolescents, and students (Fig. 3d and Fig. 4f)

We obtain the suicide rate across job status from the same sources as the main analyses^3^. We distinguish children and adolescents (individuals aged below 20), and students because some students are older than 20 years old, and some children and adolescents could be employed full-time. In 2019, the number of suicide deaths among children and adolescents was 659, while that among students was 856. These might generate some difference in the results.

##### Suicide rate across job status (Fig 4)

We obtain the suicide rate across job status from the same sources as the main analyses^3^. The results of this analyses should be interpreted with caution because job status may not be based on the information from the subjects who committed suicide. Second, to protect privacy, the cities with one or two suicide deaths (in each month) are dropped from the observations. As a result, 5.2% of the populations are dropped. Note that, because these data are collected at more aggregated level, observations are reduced from 1,848 cities to 791 cities (some cities are counted as a greater city area as a whole).

##### Municipality-level socioeconomic status (Figs. 5c and Extended Data Fig. 5c)

We use income and urban population to investigate whether the effects vary across different type of cities. We obtained these data from the “Statistical Observations of Municipalities”, published by the Ministry of Internal Affairs and Communications^4^.

##### Employment and unemployment rate (Figs. 6 a and b, Supplementary Fig. 1c, and Extended Data Fig. 5b)

The data are obtained from the Labour Force Survey from the Statistic Bureau of Japan, which is provided by the Ministry of Internal Affairs and Communications^5^.

##### Domestic violence help calls (Fig. 6c)

The data are obtained from the Gender Equality Bureau Cabinet Office. The data described are published by the 2^nd^ research meetings held by the “Research Group on the Influence of COVID-19 on Women”^6^.

##### Household income (Fig. 6d)

The data are derived from the Family Income and Expenditure Survey. The survey is conducted by the Ministry of Internal Affairs and Communications (MIAC)^7^, and the data include detailed information about household income, expenditure, and ownership of different facilities and appliances. We use total income after excluding bonus payment for our analyses.

##### Unconditional cash transfers (Fig. 6d)

The data are derived from the MIAC^8^. All Japanese citizens received cash benefits at 100,000 yen (940USD) in response to the pandemic. Note that before 5 June, 2020, 30.2% of the cash benefits were distributed and the starting date is not available. We assume that it took 2 weeks to distribute the benefits in Fig. 6d.

##### Number of Bankruptcies (Fig. 6e)

We obtain the data from Nationwide Business Failures from Tokyo Shoko Research^9^.

##### Claims for business subsidy (Fig. 6e)

We derive the data from the Ministry of Health, Labour, and Welfare^10^. The website reports the claims for a business subsidy, the number of accepted applications, and the total amount of support determined. These are reported on a weekly basis.

##### Working hours (Fig. 6f)

The data are obtained from the Monthly Labour Survey, Ministry of Health, Labour, and Welfare^11^.

##### Consumption Index (Supplementary Fig. 1c)

We obtain the data from the Consumption Activity Index, which is provided by the Bank of Japan^12^. We use the real value of the consumption index after being adjusted for inflation.

##### Diffusion Index (Supplementary Fig. 1c)

We obtain the data from the Indexes of Business Conditions, which is provided by the Cabinet Office^13^. We use the CI (Composite Indexes), measured as the Coincident Index.

##### Google Mobility Index (Supplementary Fig. 1d and Extended Data Fig. 5a)

This index is published by Google by using logs of their services (e.g. Google Maps) and describes time-series trends of geographical mobility of people in a community^*14*^.

##### COVID-19 cases in the world (Supplementary Fig. 3a)

We obtain the data from the European Centre for Disease Prevention and Control^15^, which is an agency of the European Union. The dataset includes the daily confirmed cases of COVID-19 from different countries and regions and the population. Using these two variables, we create confirmed cases per population in each country.

##### Stringency Index (Supplementary Fig. 3b)

The data are obtained from the Coronavirus Government Response Tracker, provided by the University of Oxford^16^. We use the variable “Stringency Index” that reflects seven policy responses, including school closing, workplace closing, cancel public events, restrictions on gatherings, close public transport, stay at home requirements, and restrictions on internal movement.

##### Fiscal support (Supplementary Fig. 3c)

We obtain the data from the Database of Fiscal Policy Responses to COVID-19 provided by the International Monetary Fund^17^. This variable reflects the critical fiscal measures that were announced or taken by each government in response to the COVID-19 outbreak as of June 12, 2020.

##### Weather variables

For a robustness check, we include temperature, temperature squared, precipitation, and precipitation squared as the control variables. The data are obtained from the Meteorological Agency of Japan^18^. We use weather data from 909 stations, which do not have any missing values on temperature and precipitation during our study period. Using the six monitoring stations that are closest to the population centre in each city, and the inverse of the distance from the population centre as the weights, we aggregate the data from station-level to city-level. The weights are inversely proportional to squared distance so that closer stations are given higher weights.

##### Population

We use the population to construct suicide rates and to weight all the regressions. We derive the data from the Population Estimates, managed by the Ministry of Internal Affairs and Communications (MIAC)^19^. We use population measured in 2018 across gender and age cohorts (children and adolescents: age 0∼14, adults: age 15∼64, and elderly: age over 65). Because of data limitations, the age classification is slightly different from the suicide data.

#### Supplementary Note 2. Outbreaks of COVID-19 and response of the government in Japan

Until October 2020, Japan had faced two large COVID-19 outbreaks, the first mainly from February to June 2020, and the second from July 2020 and afterward. The first case of the 2019 novel coronavirus infection (COVID-19) was confirmed in January 2020 (Supplementary Fig. 1a). By February, the number of cases was increasing rapidly. Notably, the Diamond Princess cruise ship, which docked off Yokohama Bay, was hit by the virus outbreak, with about 20% of passengers and crew infected, raising concern about the disease. In response, the national government banned large-scale gatherings and requested nationwide school closure (elementary to high school) at the end of February.

In March, despite the government’s measures, the virus spread did not slow down. Cases grew exponentially; for example, the total confirmed number of cases rose from 2,305 on April 1 to 10,283 on April 18. Given this, during this period, the government escalated anti-contagion policies. On April 7, it declared a state of emergency in seven cities that lasted about a month, and expanded it to all cities on April 16, in which citizens were requested to close schools (elementary to high school, and some universities delayed the start of their academic calendar), shops, and enterprises, and to stay home if possible. While there was high compliance, these actions were not mandated but requested (Fig. 1c, Supplementary Fig.1b). These nationwide efforts led to the containment of the virus. By the end of the May, in most prefectures, the daily new cases were brought down to below ten. Therefore, the state of emergency was lifted from prefectures where the infection situation was relatively mild on May 14, 2020, and nationwide on May 25, 2020.

However, another outbreak of COVID-19, which was larger than the first one, took place in the country. The number of infections began increasing again in late June 2020 and reached a peak in early August 2020 (around 1,500 confirmed cases per day). Unlike the large-scale health interventions implemented during the first outbreak, the national government was reluctant to enforce such extensive measures. Instead, the prefectural governments initiated tailored and focused health interventions to control disease hotspots. For example, the Tokyo prefectures requested that bars and restaurants shorten their business hours at the end of August, which led to a declining trend of the disease spread.

As is the case with many other countries, containment measures brought about an economic downturn: OECD estimated Japan’s real GDP growth declined by 6.0% with the single-hit scenario and by 7.3% with the double-hit scenario.^20^ Unemployment rates have been gradually increasing from 2.3% in January 2020 to 3.1% in October 2020.^5^ The Coincident Index (one of the indexes of business conditions) has been worsening from 94.1 in January 2020 (2015=100) to 89.7 in October 2020 (Supplementary Fig. 1c). ^13^

In response to the financial crisis, the government of Japan has made various subsidies and benefits in cash and in kind available to all citizens and those who have lost their income. These include cash benefits (100,000 yen ≈ 940 USD) for all citizens; compensation for those who have lost their income for living funds and housing, and for sick leaves; increased child allowance and support for using childcare services; monetary support including the postponement of payment for social insurance; and loans without interest and security for freelance workers^21^. Among those subsidies and benefits, the Short-Time Compensation, provided for vulnerable enterprises to stabilise their employment, plays a significant role in curbing a rise in the unemployment rate. The number of claims for business subsidy drastically increased to about 1,800,000 in late October (Fig. 6).

In our analyses, we define the pandemic period as between February 2020 and October 2020, because we believe the outbreaks in Japan and the cruise ship Diamond Princess, and the subsequent government measures, could have affected the suicidal behaviours among Japanese citizens. We further classify the period between February and June as the first outbreak, and thereafter as the second outbreak. Additionally, we define the period of the state of emergency as April and May in 2020, and the school closure (elementary to high school) as March and April 2020.

#### Supplementary Note 3. Suicide in Japan

##### Overall trend

Suicide in Japan is a serious social issue, being among the top 10 causes of death for those aged between 10 and 69: notably, it is the leading cause of death among the population aged 15-39, the second for ages 40-49, the third for ages 50-54, and the fourth for ages 55-59^22^. Additionally, suicide rates in Japan have been higher than in many other high-income countries: The Japanese suicide rate is the seventh highest among high-income countries as classified by the World Bank^23^ and OECD.^24^

##### Demography

From November 2016 to October 2020, there were 76,626 suicides with an average monthly rate of 12.8 per million population. There are large gender and age disparities in suicide rates. The suicide rate among males is about 2.3 times higher than that among females (per-month suicide rate is 18.0 per million for males and 7.8 per million for females). Particularly, males aged 20-69, who are mainly the working population, are at the highest risk of suicide (21.5 per million). A previous finding suggests that suicide deaths among males can largely be attributable to financial and economic issues. The study finds that, during the economic crisis in the 1990s, the suicide rate in Japan drastically increased, and it remained high thereafter.^25^

Although women’s suicide rate is much lower than men’s, it is the third-highest rate among OECD countries.^24^ A study investigating 193 suicide attempts suggests that family problems or loneliness could trigger mental disorders among females, while economic issues and work-related problems are the main sources among males^26^.

Suicide is one of the top causes of death among children and youth, even though those aged below 20 were at the lowest risk within the Japanese population (3.2 per million). The main causes of suicide in young generations were related to school issues. In fact, studies suggest that suicides among children and youth increase when a school session begins after a long vacation.^27^ Furthermore, suicide attempts were associated with school bullying, being a sexual minority, and juvenile delinquency.^28^

##### Social determinants

While the reasons behind suicide are extremely complex, Japan’s suicide rates have been associated with macroeconomic conditions, particularly the unemployment rate^29^, and the association is stronger than in other OECD countries^30^. During the financial crisis from 1997 to 1998, major financial institutions went bankrupt, and suicide rates sharply increased. They remained high in the next decade, when the country’s economy stagnated. In the 2010s, the unemployment rate started to decline (from 4.3% in 2012 to 2.4% in 2019), and the total number of suicides also began declining. This trend continued until recently: we observe that, on average, the suicide rate declined by 6.4% during our study period (per-month suicide rate is 13.22 per million in 2017 and 12.38 in 2019).

Although economic recession could increase suicide risk, economic policies can mitigate its negative impact. A previous finding suggests that larger government expenditures were associated with a decreased suicide rate among men aged 40-64, particularly when the unemployment rate was high^31^.

Other social factors, such as occupation, overwork^32^, marital status, and social support^33^, are reported as factors associated with suicide among the Japanese population.

##### Means

People took their own lives mainly at home, and the most common way of committing suicide was hanging, which comprised more than 60% of the cases and is a common method in many other countries^34^. Jumping (from a building or in front of a moving train) and charcoal-burning suicide (death by carbon monoxide poisoning) were also prevailing methods (around 10%).

Japan is remarkably different from the U.S. in its major suicide means. Thus, the concern in the U.S. that suicide deaths by firearm may increase during the COVID-19 pandemic subsequent to heightened gun sales^35,36^ does not appear to be relevant to Japan.

#### Supplementary Note 4. Psychological response after natural disasters

We have found that suicide declined during the first COVID-19 outbreak. This may be because of positive psychological responses to the crisis, called the pulling-together effect or honeymoon phase. Existing studies often find a drop in suicide rates after national disasters, including Hurricane Katrina and the 9/11 terrorist attack. During the COVID-19 outbreak, virus containment measures required citizens to reduce their social contacts collaboratively, and this might have generated a sense of partnership within communities. Another explanation could be that the pandemic might have altered individuals’ views on life. Facing real mortal risk, people might have started thinking of life as more precious and death as more frightening, which could have prevented them from committing suicide.

The existing literature suggests that there are six phases of a disaster^37^: (1) a Pre-Disaster phase: feelings of vulnerability, worry about safety, and responsibility for subsequent negative consequences to loved ones or damage to property; (2) the Impact phase: feelings of disbelief, numbness, fear, confusion, and panic immediately after an event; (3) the Heroic phase: predominant rescue and survival behaviours for adaptation to the environment following assistance from others; (4) the Honeymoon phase: feelings of hopefulness and optimism due to community bonding arising from sharing the catastrophic experience in addition to mutual cooperation; (5) the Disillusionment phase: feelings of disappointment and resentment due to unmet emotional and physical needs because of withdrawn supports; (6) the Reconstruction phase: during the process of attempting to revive environments, some people may become resilient and strong, while heterogeneous and intertemporal responses among individuals tend to correspond to trauma level, available resources, and coping skills.

In cases of natural disasters, a systematic review^38^ of the literature suggests that some studies report that suicide rates dropped in the initial post-disaster period (phase 4), while it eventually increased in the later period (phase 6). Although the COVID-19 pandemic and past natural disasters are distinctive in magnitude, our results showing a reduction and subsequent increase in the suicide rate are consistent with the prevalent patterns in previous literature (phases 4 and 5).

#### Supplementary Note 5. Augmented Interrupted Analysis and Different Specifications

The interrupted time-series analysis is an approach to assess the effect of large-scale intervention using longitudinal data. Because the impact of the COVID-19 pandemic is extensive, and it hit the entire country, the use of this approach seems to be reasonable. However, we think this approach could generate biased estimates for at least two reasons.

First, as discussed in the Methods section, the pandemic’s effects on the suicide rate could be confounded by seasonality. We observe that the suicide rate in February (13.0 per million) is 5.1% higher than that in January (12.4 per million) before the pandemic (2017∼2019). If we adopt interrupted time-series analysis, which compares the suicide trend between 2020 January and 2020 February (before and after the first wave of the COVID-19 pandemic), the estimates might capture the seasonal variation.

Second, the interrupted time-series analysis assumes that 1) the suicide rate changes immediately after the initial outbreak, and 2) thereafter, the slope of the time-trend is linearly altered. However, the suicide effect of the pandemic seemingly appears in different ways. During the first outbreak, the number of confirmed cases gradually increased, and people may have incrementally realised the adverse impacts of the pandemic, making the first assumption unrealistic. Additionally, the extent of the adverse effects could vary across periods during the outbreak. For instance, people may feel more stress when there are more cases, the government implements strict regulations, and economic downturn is more severe, making the second assumption unsuitable.

Here, we propose an alternative strategy to estimate the effects of the pandemic, adopting a similar framework to the interrupted analysis. To do so, we first “residualise” the suicide rates by controlling for the city-specific time trend and seasonality^39^. Intuitively, we use the “left-over” that cannot be explained by the location-specific time trend and seasonality, rather than the real suicide rate. This can be written as:

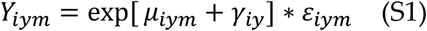

where *μ*_*iym*_ denotes the city-specific time-trend and *γ*_*iy*_ represents the city-specific month fixed effects. The residual (*e*_*iym*_) is computed as the difference between the fitted value (counterfactual) and realised value (Supplementary Fig. 4a).

Using the residual, we can try the interrupted analysis. Note that we keep the panel structure of the data instead of the longitudinal design.

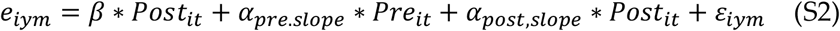

We keep the data before and after the 9 months of the COVID-19 outbreak (May 2019 to October 2020). *Pre*_*it*_ takes 1 if it is before the pandemic (May 2019 to January 2020), and *Post*_*it*_ takes 1 if it is during the pandemic (February 2020 to October 2020). *β* measures the immediate effects and the *α*_*pre*.*slope*_ and *α*_*post,slope*_ represents the linear slope of the time trend during the pre-intervention and the post-intervention.

The results show that the suicide rate declined by 2.52 points (19.8%) immediately after the intervention, and the slope becomes 0.58 points (4.6%) from −0.002 points (Supplementary Fig. 4b). However, again, it may be challenging to interpret these coefficients because the effects cannot be expressed as immediate effects (*β*) and time trend effects (*α*_*post,slope*_ -*α*_*pre*.*slope*_).

Alternatively, we estimate

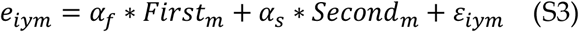

where, *First*_*m*_ and *Second*_*m*_ are analogous to equation (2). In this specification, we non-linearly measure the relative level of the residual compared to the pre-pandemic periods. We find that the suicide rate declined by 0.96 points (7.56%) during the first outbreak and increased by 2.09 points (16.4%) during the second outbreak (Supplementary Fig. 4c). These results are quantitively similar to our main results.

### Supplementary Figures

**Supplementary Fig. 1.**
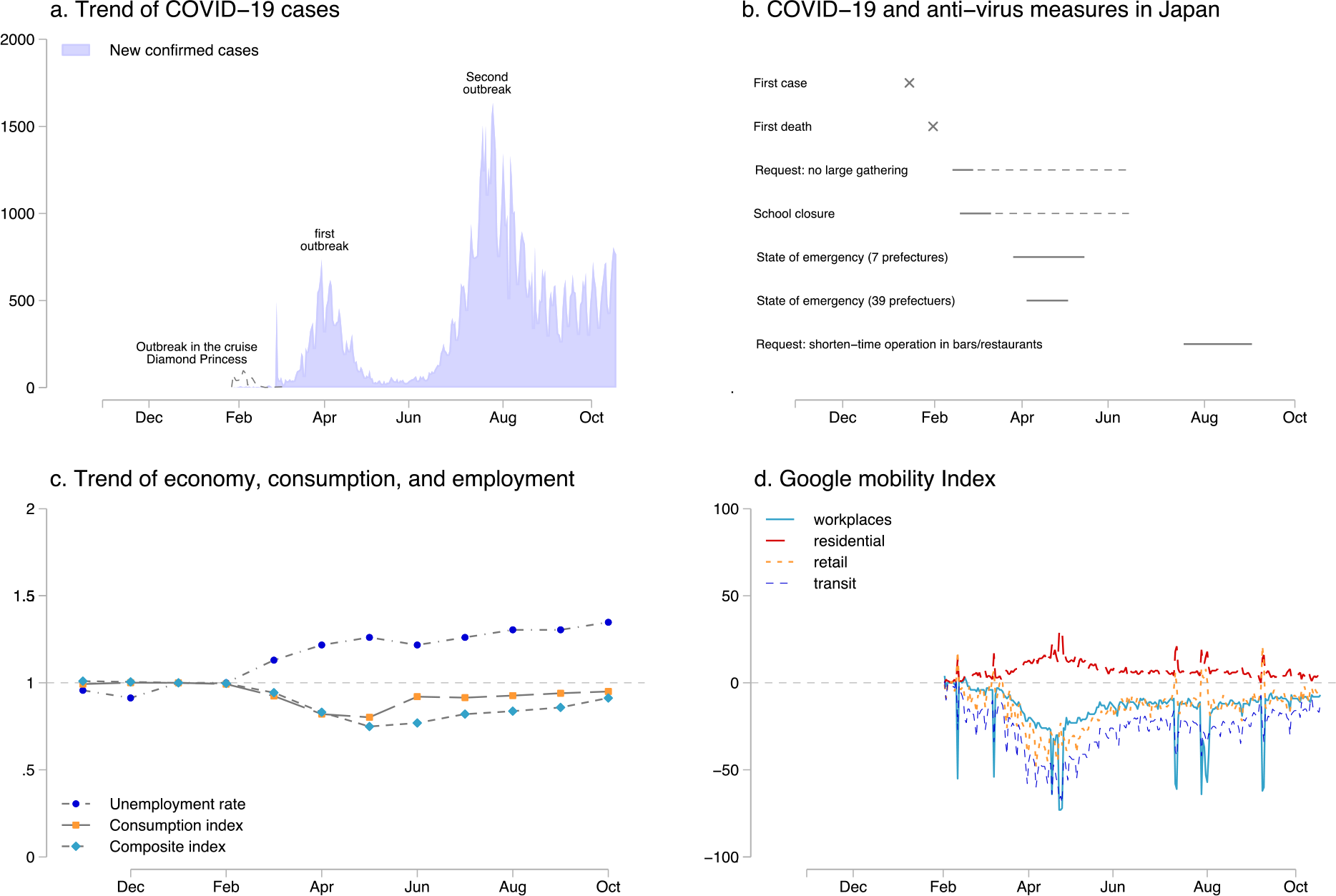
COVID-19, the virus containment measures, macroeconomic conditions, and human movement: **Panel a** describes daily confirmed new COVID-19 cases. We also present the outbreak on the cruise ship Diamond Princess because it docked off Yokohama Bay. **Panel b** represents the major dates of the pandemic outbreak and the government’s response to the disease outbreak. **Panel c** shows the macroeconomic conditions, including unemployment rate, consumption index, and the composite index. (Jan 2020 = 1.00). **Panel d** draws the Google mobility index relative to 15 February 2020.

**Supplementary Fig. 2.**
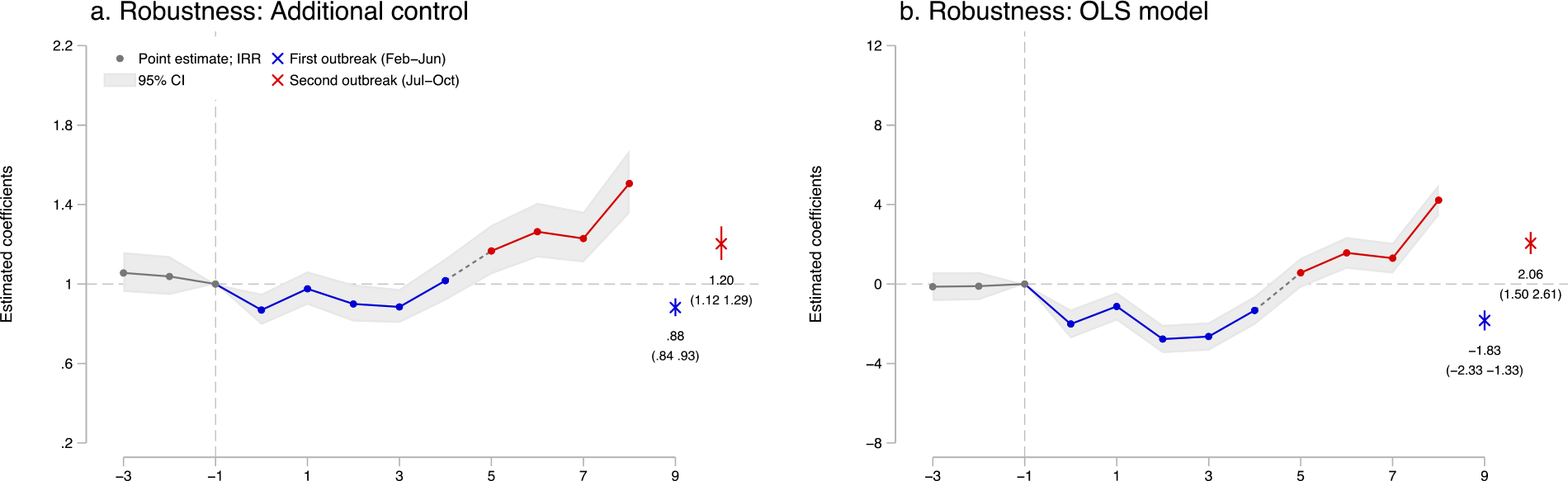
Robustness check: **Panel a** describes the results when including the time-varying controls and prefecture-specific quadratic trend. The time-varying controls include monthly average temperature and precipitation and their squares. The results are comparable. **Panel b** uses the suicide rate and the OLS instead of the Poisson model, which shows the similar trend of suicide rate. All regressions include city-by-year fixed effects and city-by-month fixed effects and are weighted by population, and standard errors are clustered at the city level. N= 61,209 (a), and 88,512 (b). The separated observations are dropped for Poisson model regression (see Methods)

**Supplementary Fig. 3.**
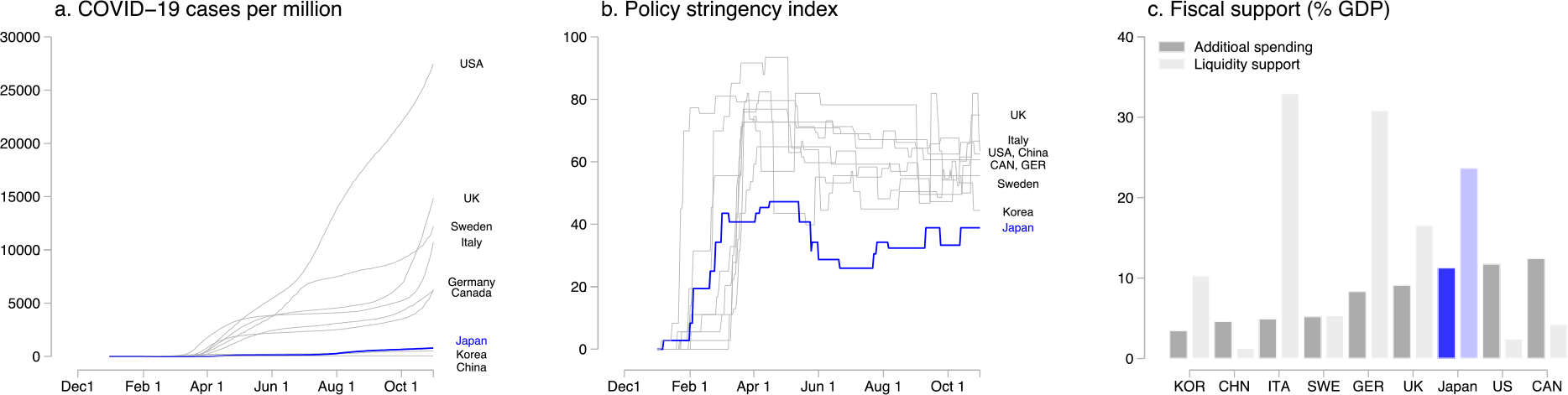
International comparison of COVID-19 cases, health interventions, and fiscal policies: **Panels a, b, and c** document the confirmed COVID-19 cases per population in different countries (panel a), the stringency index of the virus containment health intervention (b), and fiscal support as a percentage share of GDP (additional spending and liquidity support) (c). The data sources are described in Supplementary Note 1.

**Supplementary Fig. 4.**
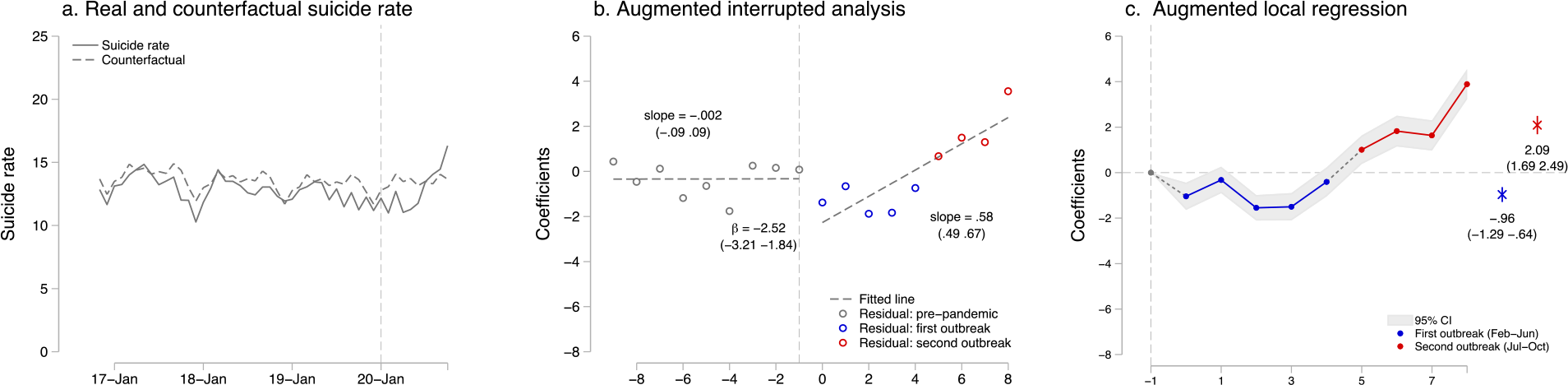
Robustness checks and time interrupted analysis: **Panel a** represents real suicide rate and predicted suicide rate estimated by the city-specific time-trend and the city-specific month fixed effects (Counterfactual). **Panel b** shows the result of the augmented interrupted analysis by using the residual, assuming that the pandemic effects are revealed as the immediate impact and slope change. **Panel c** represents relative levels of the residual compared to the pre-pandemic periods, distinguishing between the first and second outbreaks.

### Supplementary Tables

**Supplementary Table 1.** Summary statistics.

| Suicide rate<br>(per million) | All Periods<br>(Nov 2016-Oct 2020) | Before the<br>Pandemic<br>(Nov 2016-Jan 2020) | First<br>Outbreak<br>(Feb-Jun 2020) | Second<br>Outbreak<br>(Jul-Oct 2020) |
| --- | --- | --- | --- | --- |
| All | 12.7<br>(17.1) | 12.8<br>(17.1) | 11.6<br>(15.1) | 14.6<br>(19.3) |
| *Panel a: by gender* |  |  |  |  |
| Male | 18.0<br>(33.7) | 18.1<br>(34.5) | 16.4<br>(26.5) | 18.9<br>(31.4) |
| Female | 7.8<br>(19.2) | 7.8<br>(19.8) | 7.0<br>(14.9) | 10.5<br>(17.8) |
| *Panel b: by age* |  |  |  |  |
| Age below 20 | 3.2<br>(21.6) | 3.1<br>(20.4) | 3.2<br>(22.8) | 4.8<br>(30.0) |
| Age 20-69 | 15.2<br>(25.4) | 15.4<br>(25.0) | 13.5<br>(22.7) | 17.0<br>(31.5) |
| Age over 70 | 12.1<br>(27.8) | 12.1<br>(28.1) | 11.5<br>(25.1) | 14.0<br>(27.5) |
| *Panel c: by job status* |  |  |  |  |
| Self-employed | 0.45<br>(1.69) | 0.46<br>(1.73) | 0.43<br>(1.61) | 0.4<br>(1.5) |
| Employed | 2.30<br>(4.24) | 2.31<br>(4.26) | 2.03<br>(3.92) | 3.0<br>(4.8) |
| Unemployed | 1.85<br>(3.65) | 1.88<br>(3.71) | 1.69<br>(3.18) | 2.0<br>(3.7) |
| Housewife | 0.43<br>(1.39) | 0.43<br>(1.38) | 0.36<br>(1.30) | 0.6<br>(1.8) |
| Retired | 1.91<br>(3.96) | 1.96<br>(4.02) | 1.65<br>(3.60) | 2.1<br>(4.0) |

*Notes:* This table shows the mean and standard deviation (in the brackets) of each variable. The results are weighted by population. We include 1,849 cities. For gender and age groups, we use the cohort-specific population to compute the suicide rate, while we use the total population for suicide rate across different job status.

**Supplementary Table 2.**
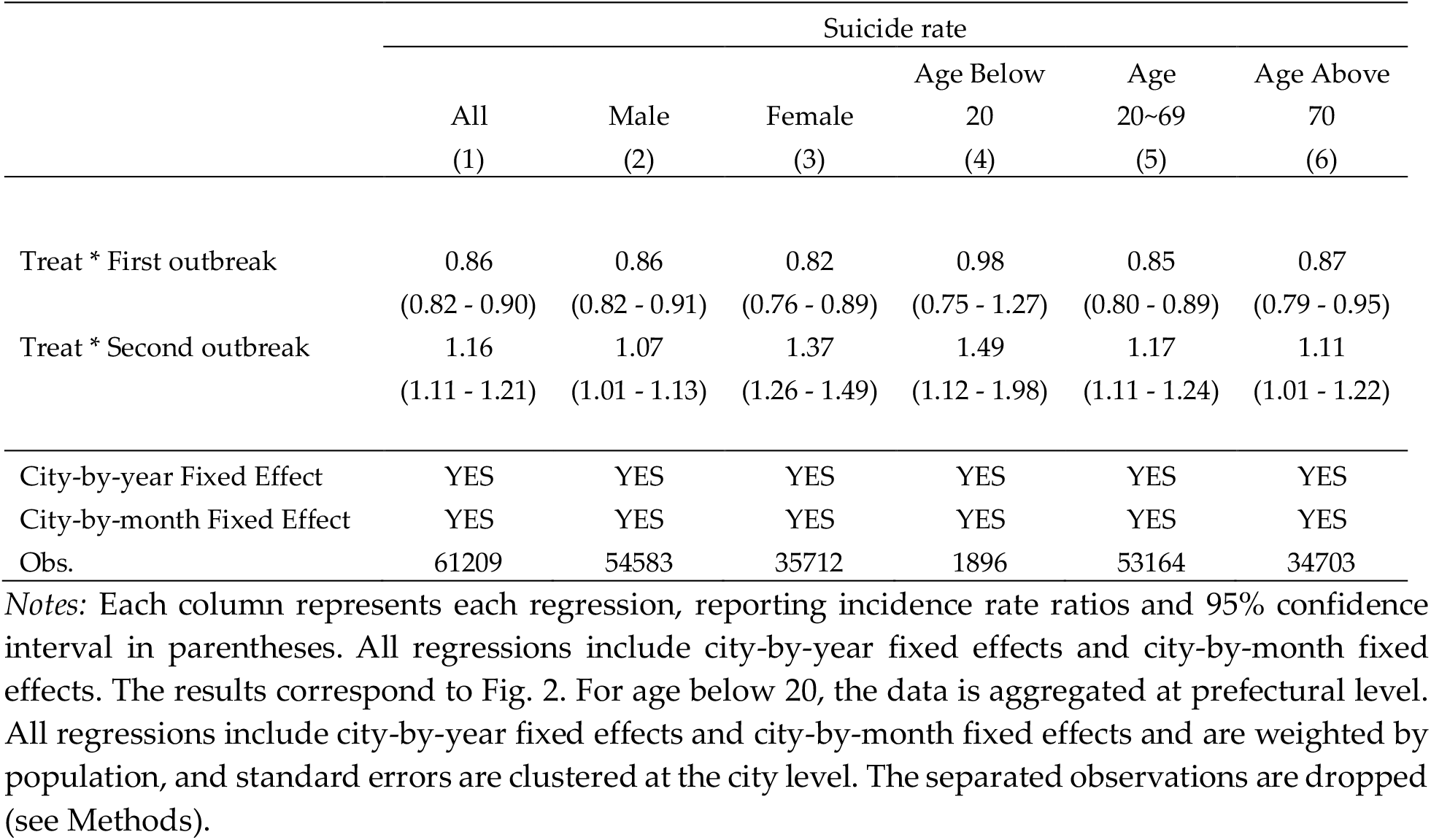
Full results of the effects of COVID-19 pandemic on suicide across gender and age groups using DID.

**Supplementary Table 3.** Full results of the effects of COVID-19 pandemic on suicide across gender and age groups in each month using DID.

|  | Suicide rate |  |  |  |  |  |
| --- | --- | --- | --- | --- | --- | --- |
|  | All | Male | Female | Age Below 20 | Age 20~69 | Age Above 70 |
|  | (1) | (2) | (3) | (4) | (5) | (6) |
| Treat * Feb | 0.84<br>(0.78 - 0.91) | 0.87<br>(0.80 - 0.95) | 0.77<br>(0.67 - 0.89) | 0.90<br>(0.59 - 1.35) | 0.84<br>(0.77 - 0.92) | 0.86<br>(0.72 - 1.02) |
| Treat * Mar | 0.92<br>(0.86 - 0.98) | 0.92<br>(0.85 - 1.00) | 0.90<br>(0.80 - 1.02) | 1.11<br>(0.74 - 1.66) | 0.93<br>(0.86 - 1.01) | 0.84<br>(0.73 - 0.97) |
| Treat * Apr | 0.80<br>(0.75 - 0.86) | 0.82<br>(0.76 - 0.90) | 0.74<br>(0.64 - 0.84) | 0.77<br>(0.54 - 1.08) | 0.79<br>(0.72 - 0.86) | 0.85<br>(0.73 - 0.98) |
| Treat * May | 0.81<br>(0.76 - 0.87) | 0.81<br>(0.74 - 0.88) | 0.78<br>(0.69 - 0.89) | 0.90<br>(0.56 - 1.43) | 0.78<br>(0.72 - 0.85) | 0.88<br>(0.76 - 1.01) |
| Treat * Jun | 0.91<br>(0.85 - 0.98) | 0.89<br>(0.81 - 0.97) | 0.93<br>(0.81 - 1.05) | 1.24<br>(0.85 - 1.82) | 0.89<br>(0.81 - 0.97) | 0.91<br>(0.79 - 1.05) |
| Treat * Jul | 1.05<br>(0.98 - 1.13) | 0.99<br>(0.91 - 1.07) | 1.18<br>(1.05 - 1.33) | 1.17<br>(0.74 - 1.85) | 1.03<br>(0.95 - 1.12) | 1.08<br>(0.94 - 1.24) |
| Treat * Aug | 1.14<br>(1.06 - 1.22) | 1.08<br>(1.00 - 1.18) | 1.27<br>(1.12 - 1.44) | 1.77<br>(1.21 - 2.60) | 1.14<br>(1.05 - 1.24) | 1.05<br>(0.91 - 1.20) |
| Treat * Sep | 1.10<br>(1.03 - 1.18) | 1.04<br>(0.95 - 1.12) | 1.26<br>(1.12 - 1.42) | 1.49<br>(1.11 - 2.00) | 1.11<br>(1.03 - 1.21) | 1.05<br>(0.91 - 1.22) |
| Treat * Oct | 1.36<br>(1.28 - 1.45) | 1.18<br>(1.09 - 1.28) | 1.82<br>(1.62 - 2.04) | 1.50<br>(1.00 - 2.25) | 1.42<br>(1.32 - 1.53) | 1.28<br>(1.11 - 1.47) |
| City-by-year<br>Fixed Effect | YES | YES | YES | YES | YES | YES |
| City-by-month<br>Fixed Effect | YES | YES | YES | YES | YES | YES |
| Obs. | 61209 | 54583 | 35712 | 1896 | 53164 | 34703 |

*Notes:* Each column represents each regression, reporting incidence rate ratios and 95% confidence intervals in parentheses. For age below 20, the data is aggregated at prefectural level. All regressions include city-by-year fixed effects and city-by-month fixed effects and are weighted by population, and standard errors are clustered at the city level. The separated observations are dropped (see Methods).

**Supplementary Table 4.** Full results of the effects of COVID-19 pandemic on suicide across gender and age groups using DID.

|  | Suicide rate |  |  |  |  |  |
| --- | --- | --- | --- | --- | --- | --- |
|  | All | Male | Female | Age<br>Below 20 | Age<br>20~69 | Age Above<br>70 |
|  | (1) | (2) | (3) | (4) | (5) | (6) |
| FD2 | 1.02<br>(0.94 - 1.11) | 1.01<br>(0.90 - 1.12) | 1.05<br>(0.89 - 1.24) | 1.01<br>(0.63 - 1.63) | 1.05<br>(0.95 - 1.17) | 0.94<br>(0.79 - 1.12) |
| FD1 | 1.01<br>(0.92 - 1.10) | 1.00<br>(0.90 - 1.12) | 1.03<br>(0.88 - 1.22) | 0.89<br>(0.59 - 1.34) | 1.04<br>(0.94 - 1.15) | 0.95<br>(0.78 - 1.15) |
| D1 | 0.85<br>(0.78 - 0.93) | 0.87<br>(0.78 - 0.97) | 0.79<br>(0.67 - 0.94) | 0.87<br>(0.55 - 1.37) | 0.87<br>(0.78 - 0.96) | 0.82<br>(0.67 - 1.01) |
| D2 | 0.93<br>(0.86 - 1.01) | 0.93<br>(0.84 - 1.02) | 0.93<br>(0.80 - 1.08) | 1.08<br>(0.69 - 1.67) | 0.96<br>(0.87 - 1.06) | 0.81<br>(0.68 - 0.97) |
| D3 | 0.81<br>(0.75 - 0.88) | 0.83<br>(0.74 - 0.91) | 0.76<br>(0.65 - 0.89) | 0.74<br>(0.50 - 1.11) | 0.81<br>(0.73 - 0.90) | 0.82<br>(0.68 - 0.97) |
| D4 | 0.82<br>(0.75 - 0.89) | 0.81<br>(0.73 - 0.90) | 0.80<br>(0.69 - 0.93) | 0.87<br>(0.54 - 1.41) | 0.80<br>(0.73 - 0.89) | 0.84<br>(0.71 - 1.00) |
| D5 | 0.92<br>(0.84 - 1.01) | 0.89<br>(0.80 - 1.00) | 0.95<br>(0.81 - 1.11) | 1.21<br>(0.78 - 1.88) | 0.92<br>(0.82 - 1.02) | 0.87<br>(0.74 - 1.04) |
| D6 | 1.06<br>(0.98 - 1.15) | 0.99<br>(0.90 - 1.10) | 1.22<br>(1.05 - 1.41) | 1.14<br>(0.72 - 1.80) | 1.06<br>(0.96 - 1.17) | 1.04<br>(0.88 - 1.24) |
| D7 | 1.15<br>(1.05 - 1.25) | 1.09<br>(0.98 - 1.21) | 1.30<br>(1.12 - 1.52) | 1.72<br>(1.16 - 2.55) | 1.18<br>(1.06 - 1.31) | 1.01<br>(0.85 - 1.20) |
| D8 | 1.11<br>(1.02 - 1.21) | 1.04<br>(0.94 - 1.15) | 1.29<br>(1.11 - 1.50) | 1.45<br>(1.06 - 1.98) | 1.15<br>(1.04 - 1.27) | 1.01<br>(0.85 - 1.21) |
| D9 | 1.38<br>(1.27 - 1.49) | 1.18<br>(1.07 - 1.31) | 1.87<br>(1.61 - 2.16) | 1.46<br>(0.94 - 2.27) | 1.46<br>(1.33 - 1.61) | 1.23<br>(1.04 - 1.46) |
| City-by-year Fixed<br>Effect | YES | YES | YES | YES | YES | YES |
| City-by-month<br>Fixed Effect | YES | YES | YES | YES | YES | YES |
| Obs. | 61209 | 54583 | 35712 | 1896 | 53164 | 34703 |

*Notes:* Each column represents each regression, reporting incidence rate ratios and 95% confidence intervals in parentheses. All regressions include city-by-year fixed effects and city-by-month fixed effects. The results correspond to Fig. 2. All regressions include city-by-year fixed effects and city-by-month fixed effects and are weighted by population, and standard errors are clustered at the city level. The separated observations are dropped (see Methods).

**Supplementary Table 5.** Full results of the heterogeneous effects of the COVID-19 pandemic among age groups and gender, before and after the state of emergency and the school closure.

| Dependent variable: | All | Male adult | Female adult | Male elderly | Female elderly | Age Below 20 |
| --- | --- | --- | --- | --- | --- | --- |
| Suicide rate | (1) | (2) | (3) | (4) | (5) | (6) |
| Treat * COVID-19 outbreak<br>(baseline in Fig 4) | 0.99<br>(0.95 - 1.03) |  |  |  |  |  |
| Treat * Statement of Emergency<br>(Panel a in Fig 4) |  | 0.79<br>(0.73 - 0.85) | 0.73<br>(0.65 - 0.83) | 0.85<br>(0.73 - 1.00) | 0.80<br>(0.64 - 1.00) |  |
| Treat * First outbreak, other period<br>(Panel b in Fig 4) |  | 0.9<br>(0.83 - 0.96) | 0.85<br>(0.76 - 0.95) | 0.87<br>(0.75 - 1.00) | 0.82<br>(0.67 - 1.00) |  |
| Treat * Second outbreak<br>(Panel c in Fig 4) |  | 1.07<br>(1.00 - 1.14) | 1.44<br>(1.30 - 1.60) | 1.03<br>(0.90 - 1.18) | 1.27<br>(1.08 - 1.51) |  |
| Treat * Before school closure<br>(Panel d in Fig 4) |  |  |  |  |  | 0.90<br>(0.60 - 1.36) |
| Treat * During school closure<br>(Panel d in Fig 4) |  |  |  |  |  | 0.91<br>(0.66 - 1.26) |
| Treat * After school closure<br>(Panel d in Fig 4) |  |  |  |  |  | 1.35<br>(1.04 - 1.75) |
| City-by-year Fixed Effect | YES | YES | YES | YES | YES | YES |
| City-by-month Fixed Effect | YES | YES | YES | YES | YES | YES |
| Obs. | 61209 | 47317 | 26319 | 24478 | 16531 | 1896 |

*Notes:* Each column represents each regression, reporting incidence rate ratios and 95% confidence intervals in parentheses. All regressions include city-by-year fixed effects and city-by-month fixed effects. The results correspond to Fig. 3. For age below 20, the data is aggregated at prefectural level. All regressions include city-by-year fixed effects and city-by-month fixed effects and are weighted by population, and standard errors are clustered at the city level. The separated observations are dropped (see Methods).

**Supplementary Table 6.** Full results of heterogeneous effects of the COVID-19 pandemic across job status.

| Dependent variable: | Employed<br>(Panel a) | Retired<br>(Panel b) | Unemployed<br>(Panel c) | Self-employed<br>(Panel d) | Housewife<br>(Panel e) | Students<br>(Panel f) |
| --- | --- | --- | --- | --- | --- | --- |
| Suicide rate | (1) | (2) | (3) | (4) | (5) | (6) |
| Treat * First outbreak | 0.80<br>(0.70 - 0.91) | 0.80<br>(0.69 - 0.92) | 0.83<br>(0.73 - 0.95) | 0.81<br>(0.61 - 1.09) | 1.17<br>(0.81 - 1.70) |  |
| Treat * Second outbreak | 1.24<br>(1.11 - 1.40) | 1.08<br>(0.94 - 1.24) | 1.11<br>(0.95 - 1.29) | 0.95<br>(0.68 - 1.33) | 2.32<br>(1.65 - 3.26) |  |
| Treat *<br>Before school closure |  |  |  |  |  | 0.98<br>(0.61 - 1.57) |
| Treat *<br>School closure |  |  |  |  |  | 0.51<br>(0.34 - 0.77) |
| Treat *<br>After school closure |  |  |  |  |  | 1.05<br>(0.77 - 1.43) |
| City-by-year Fixed Effect | YES | YES | YES | YES | YES | YES |
| City-by-month Fixed Effect | YES | YES | YES | YES | YES | YES |
| Obs. | 10723 | 9146 | 9246 | 4124 | 3854 | 3220 |

*Notes:* Each column represents each regression, reporting incidence rate ratios and 95% confidence intervals in parentheses. All regressions include city-by-year fixed effects and city-by-month fixed effects. The results correspond to Fig. 4. For age below 20, the data is aggregated at prefectural level. All regressions include city-by-year fixed effects and city-by-month fixed effects and are weighted by population, and standard errors are clustered at the city level. The separated observations are dropped (see Methods).

**Supplementary Table 7.**
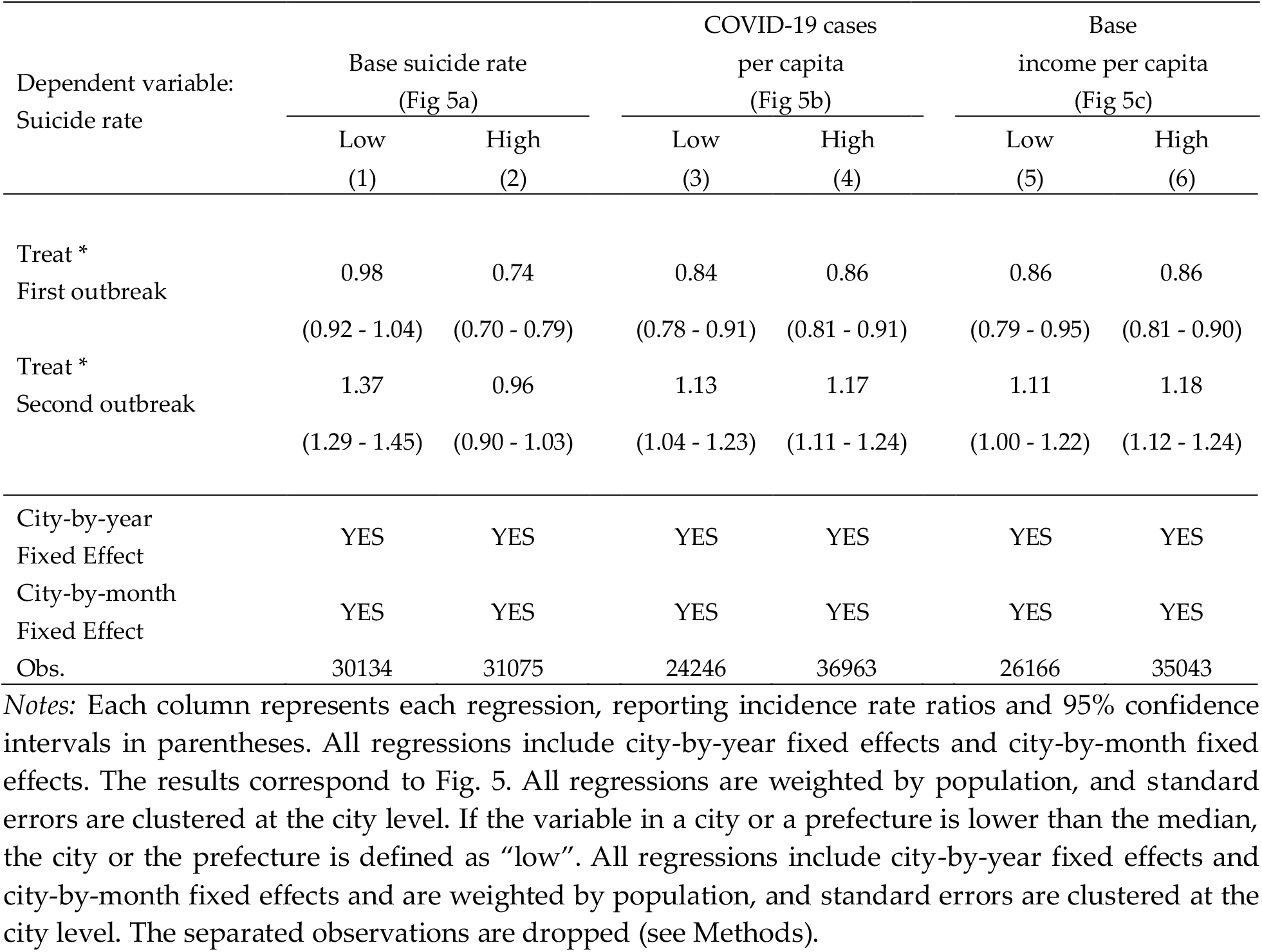
Full results of heterogeneous effects of the COVID-19 pandemic across geography.

## Notes

### Competing Interest Statement

The authors have declared no competing interest.

### Funding Statement

Shohei Okamoto is supported by the postdoctoral fellowship of the Japan Society for the Promotion of Science (No. 20J00394) and the Murata Science Foundation.

### Author Declarations

Ethical approval was not requred as this research is based on secondary analysis of publicly available data.

